# Hairpin structure facilitates multiplex high-fidelity DNA amplification in real-time PCR

**DOI:** 10.1101/2021.11.10.21266179

**Authors:** Kerou Zhang, Alessandro Pinto, Lauren Yuxuan Cheng, Ping Song, Peng Dai, Michael Wang, Luis Rodriguez, Cailin Weller, David Yu Zhang

## Abstract

Clinically and biologically, it is essential to detect rare DNA-sequence variants for early cancer diagnosis or drug-resistance mutations identification. Some of the common quantitative PCR (qPCR)-based variant detection methods are restricted in the limit of detection (LoD) because the DNA polymerases used have a high polymerase misincorporation rate, thus the detection sensitivity is sometimes unsatisfactory. With the proofreading activity, high-fidelity (HiFi) DNA polymerases have a 50- to 250-fold higher fidelity. However, there are currently no proper probe-based designs as the fluorescence indicator allowing multiplexed HiFi qPCR reactions, thus restricting the application of HiFi DNA polymerases like the variant detection. We presented the Occlusion System, composed of a 5’-overhanged primer with fluorophore modification and a probe with a short-stem hairpin and a 3’ quencher modification. We demonstrated that the Occlusion System allowed multiplexing HiFi qPCR reaction, and it was compatible with the current variant-enrichment method to improve the LoD up to 10-fold. Thus, the Occlusion System satisfactorily functioned as efficient fluorescence indicator in HiFi qPCR reactions and allowed application of HiFi DNA polymerases in variant detection methods to improve detection sensitivity.

## Introduction

The Polymerase chain reaction (PCR) has been one of the most widely used methods in bio-laboratories. It can be used in conjunction with intercalating dyes or fluorophore-labeled probes to allow quantitative and real-time detection of nucleic acid targets using a fluorescence detection device, or it can be used upstream of a high-throughput sequencing reaction for target enrichment. Compared with the high-throughput sequencing methods, quantitative PCR (qPCR) reduces the total turnaround time with maintained or increased analytical sensitivity and reproducibility, benefiting the molecular diagnostics of diseases requiring timely diagnosis and treatment with clinical sensitivity and specificity, like clinical oncology and clinical microbiology^1–9^.

Profiling nucleic acid variants with low allele frequencies, such as cancer mutations from liquid biopsy samples, drug-resistance mutations, or pathogenic mutations from different types of clinical specimens^9^, is clinically essential. Genetic alternations could result in a mixed population of mutant and normal cells with the tissue, which could progressively lead to oncogenesis^10^. Earlier diagnosis could be achieved with more sensitive detection of the low-abundance mutations for tumor indication, thus better prognosis and precision medicine^11^. Liquid biopsy has revealed an advantage over traditional gold standard tissue biopsy in its non-invasiveness, good concordance in genome profiling, and compatibility with different sample types^12,13^, thus liquid biopsy could be suitable for aiding early diagnosis^14,15^. However, somatic mutations detected from the liquid biopsy usually have lower variant allele frequency (VAF) than those from the tissue biopsy^16^, which added to the difficulty of sensitive and accurate detection of early-stage somatic mutations. Besides, liquid biopsy could offer a potential solution for longitudinal mutation profiling in diseases like minimal residual disease (MRD)^17,18^, which usually requires sensitivity down to 1:10^4^ to 1:10^6^. Similarly, the need to detect low mutation rates within the drug-resistant *Mycobacterium tuberculosis* strains^19^ or drug-resistance colorectal cancer^20^ has been reported.

Some common qPCR diagnostics methods using DNA polymerases lacking proofreading activity, like blocker displacement amplification (BDA)^21^, allele-specific PCR (As-PCR)^22,23^, amplification-refractory mutation system (ARMS-PCR)^24,25^, competitive allele-specific TaqMan PCR (Cast-PCR)^26,27^, and co-amplification at lower denaturation temperature PCR (COLD-PCR)^28^, mainly have a limit of detection (LoD) around 0.1% to 1% VAF which is no longer satisfactory^29^. The high polymerase misincorporation rate^30^ from the lack of the proofreading activity could introduce false-positive mutations into generated amplicons, thus obscuring readouts in qPCR methods and restricting the LoD (Figure S1A). As the need to detect mutations with VAF low to or even lower than 0.01% VAF has increased for both basic research and clinical diagnosis, researchers and scientists have been exploring the solutions. Scientists have reported integrating CRISPR^31^ or Argonaute-mediated^32,33^ cleavage of targets with non-HiFi PCR to achieve rare mutation detection with improved sensitivity (LoD ≤0.01% VAF). Allele-specific BDA (As-BDA)^34^ was also capable of detecting and profiling multiplexed mutations down to 0.01% VAF within the single tube using Taq DNA polymerases by implementing allele-specific TaqMan probes. However, introducing extra components or steps before or into the qPCR reaction may increase system complexity. Moreover, As-BDA relies much on the preliminary knowledge of target information, limiting its application in the discovery of new biomarkers. HiFi DNA polymerases possess the 3’ to 5’ exonuclease activity known as proofreading features^35–38^, leading to 50- to 250-fold increased fidelity^30^. Hence, HiFi DNA polymerases have the potential to improve the LoD of the previous design and achieve up to 10-fold increased sensitivity in the qPCR reaction (Figure S1B). Moreover, HiFi DNA polymerases are prevalent in biolaboratory and can be easily adapted to current qPCR-based diagnostic methods.

The fluorescence indicator is necessary to indicate amplicon generation in qPCR reactions. Besides intercalating dye, polymerases without the proofreading activity are usually compatible with multiple fluorescence-indicating methods, including molecular beacons^39^, Sunrise primers^40^, Scorpion primers^41^, hydrolysis (TaqMan) probes^42^ or fluorescence resonance energy transfer (FRET) probes^43^, to indicate the production of PCR amplicons. Probe-based tools like the TaqMan probe are generally favored over intercalating dyes because multiple different spectrally distinct fluorophores can be used on different probes simultaneously to allow multiplexed detection and quantitation of 2-6 distinct DNA target species in a single reaction. However, HiFi DNA polymerases have only been reported compatible with limited choice^44^. Designs like TaqMan probes and Sunrise primers have been demonstrated to be incompatible with HiFi DNA polymerase (Figure S2) for single target detection, let alone multiplexing. A key reason is that the 3’ to 5’ exonuclease activity of the HiFi polymerases would digest the 3’ mismatched sequence or 3’ chemical modification within the double-stranded region formed between the primer or probe and the template. Also, HiFi polymerases usually lack the 5’ to 3’ exonuclease activity to produce detectable signals with most probe designs. Thus, finding an appropriate and efficient probe-based fluorescence indicator design, which is compatible with HiFi DNA polymerases, cost-efficient, easily accessible, and plex-scalable in qPCR, has been challenging.

Here, we present the Occlusion System, composed of Occlusion Primer and Occlusion Probe, providing a straightforward solution for simultaneously detecting multiplexed targets within the HiFi qPCR reactions as a probe-based fluorescence indicator tool. Moreover, the Occlusion System, is compatible with other variant enrichment (diagnostic) methods like BDA in single-plex and multi-plex settings. In terms of analytical performance, we demonstrated that the Occlusion BDA System in HiFi qPCR reactions (HiFi BDA Occlusion System) was sensitive in VAF down to 0.01% using synthetic variants, of which the LoD was improved up to 10-fold compared with those of Taq-based BDA qPCR reactions. Furthermore, the HiFi BDA Occlusion System was accurate in VAF calling with a percentage error less than 23.1% on commercial reference samples. We further applied the HiFi BDA Occlusion System in peripheral blood mononuclear cell (PBMC) samples from 27 acute myeloid leukemia (AML) patients and 15 healthy donors, and detected 6 *FLT3* mutations, 2 *IDH1* mutations in 27 patients. Comparative results between the HiFi BDA Occlusion qPCR and the droplet digital PCR (ddPCR) in the *FLT3* mutations showed 100% concordance in mutations over 0.1% VAF with limited DNA input. In brief, the Occlusion System achieved stable fluorescence indication for both single-plex or multi-plex HiFi qPCR reactions and improved clinical detection limit by 10-fold in qPCR reactions by enabling the integration of the previous multiplexed variant-enrichment method and HiFi DNA polymerases.

## Materials and Methods

### Study samples

Twenty-seven AML PBMC samples were purchased from Discovery Life Science (DLS) with written informed patient consent. Fifteen PBMC samples from healthy volunteers were purchased from Zen-Bio, who collected the samples with written informed patient consent. IRB approval was not required as these are Exemption 4 (commercial, de-identified samples). Detailed sample information is summarized in Tables S2 and S3.

### Oligonucleotides and repository samples

Primers, BDA blockers, Occlusion primers, Occlusion probes and synthetic double-stranded DNA fragments (gBlocks) were purchased from Integrated DNA Technologies (IDT). Detailed design sequence was summarized in Tables S15 and S16. Primers, BDA blockers and gBlocks were purchased as standard desalting-purified, Occlusion primers and Occlusion probes as HPLC-purified; they were resuspended in 1× IDTE buffer (10 mM Tris, 0.1 mM EDTA) in 100 uM (gBlocks were resuspended in 10 ng/μL) as the original stock and stored at 4 °C. The dilution buffer composed of 100 ng/μL carrier RNA (Qiagen, 1017647) in 1× IDTE buffer with 0.1% TWEEN 20 (Sigma Aldrich), was used to serially dilute gBlocks. Human cell-line gDNA NA18562 and NA18537 repository samples (Coriell Biorepository), Commercial reference gDNA samples (Tru-Q 7 (1.3% Tier) Reference Standard (HD734) and Myeloid DNA Reference Standard (HD829)) (Horizon Discovery) were diluted with 1x IDTE buffer (IDT). All the dilutions and original stock were stored at 4 °C for short-term usage, −20 °C for long-term storage except Horizon reference gDNA samples (both dilutions and original stock were stored at 4 °C).

### DNA extraction from PBMC samples

DNA was extracted from PBMC samples using the DNA Blood Mini kit (Qiagen, 51104). NanoDrop spectrophotometer was used to measure the yield of DNA. All DNA materials were stored at -20 °C until ready for analysis.

### Reference material preparation

Synthetic gBlocks, used for mimicking natural mutations, were quantified by qPCR. The concentration of gBlocks were then adjusted by comparing cycle threshold (Ct) values with that of 40 ng/μL human gDNA NA18562. Then gBlocks were further diluted with gDNA NA18562 to prepare reference samples of 20%, 10%, 5%, 1%, 0.5%, 0.3%, 0.1%, 0.05%, 0.03%, 0.01% and 0.005% VAF by serial dilution. 10% and 1% VAF reference samples were quantitated in NGS before further dilution to lower-VAF samples.

### Occlusion qPCR protocol

All Occlusion qPCR assays were performed in the CFX96 Touch Real-Time PCR Detection System using 96-well plates. Phusion Hot Start Flex 2X Master Mix (Thermo Scientific) was used for each qPCR reaction. For the single-plex HiFi qPCR reactions not using the Occlusion System, the SYTO 13 green-fluorescent nucleic acid stain is used to indicate amplicon generation. Roughly 200ng DNA in 5 µL were loaded as input into each reaction as default, unless otherwise noted. The thermocycling program started with 30 s polymerase activation and initial denaturation at 98 °C, followed by 55 cycles of 10 s at 98 °C for DNA denaturing, 30 s at 63 °C for annealing and 30 s at 72 °C for extension (98 °C: 30s - (98 °C: 10 s - 63 °C: 30 s - 72 °C: 30 s) x 55). Fluorescence signal was collected at 63 °C per cycle. All the reactions were conducted in duplicate or triplicate. Details of composition concentration are summarized in Tables S12 to S14.

### ddPCR quantitation protocol

ddPCR mutation assays from Bio-Rad were used to quantitate mutations in the *FLT3* gene [dHsaMDS2514588 for *FLT3* 2503G>A (D835N), dHsaMDS420144135 for *FLT3* 2508_2510del (I836del), dHsaMDV2010047 for *FLT3* 2503G>T (D835Y), dHsaMDV2510492 for *FLT3* 2503G>C (D835H)]. The whole workflow was conducted on a Q200 Droplet Digital PCR System (Bio-Rad). A 20 µL reaction containing 10 µL 2X ddPCR Supermix for Probes (No dUTP) (Bio-Rad, 1863024), 1 µL 20x target (FAM) and wildtype (HEX) primers/probe and roughly 20ng samples was prepared and loaded into the middle wells of DG8 cartridges (Bio-Rad, 1864008). 70 µL of the Droplet Generation Oil for Probes (Bio-Rad, 1963005) was then added to the top wells of the cartridges. The DG8 cartridges were put in the QX200 Droplet Generator (Bio-Rad, 10031907) for droplet generation. Then the thermo cycling, with all ramp speed set at 2 °C/sec, started with 10 min at 95 °C, followed by 40 repeated cycles of 30 s at 94 °C and 1 min at 55 °C. There is a 10 min incubation step at 98 °C following the repeated cycle before holding the plate at 4 °C until the next step. The plate was then loaded onto the QX200 Droplet Reader (Bio-Rad, 1864003) for droplet counts. QuantaSoft software was used to collect and analyze the data. Further data analysis on MATLAB used the code published previously^21^.

## Results

### Hairpin-based Fluorescence indicator in HiFi qPCR reactions

To find a probe-based design as the fluorescence indicator in multiplexed HiFi qPCR reactions, we previously attached a fluorophore-modified overhang sequence to the 5’ region of the conventional primer, reverse complementary to a companion strand modified with a 3’ quencher. However, the quencher was digested by the proofreading feature of HiFi DNA polymerases during qPCR, resulting in an abnormal amplification curve from the initial cycle at the absence of DNA templates (Figure 1A). Inspired by the functionality of RNA hairpins in intrinsic transcription termination^45–48^, where the polymerase would fall off and end the transcription once encountering the hairpin termination structure, we hypothesized that hairpin structure could efficiently hinder the proofreading feature of HiFi DNA polymerases. Then we experimentally found that the HiFi DNA polymerases could still generate abnormally increased fluorescence signal per cycle when the hairpin structure attached to the 3’ region of the companion strand (Figure 1B). As known, a 10 to 20nt priming region is enough for the initiation of nucleic acid synthesis^49^. It was suspicious that the HiFi DNA polymerases could still bind to the stem of the hairpin (20nt in Figure 1B) on the companion strand, digest the 3’ quencher modification and extend on the companion strand. The 5’ fluorophore modification attached to the primer may destabilize the hybridization of the 5’ overhang region and the companion strand, thus increasing the chance of strand dissociation^50^, leading to the extension of the long-stem region of the hairpin on the companion strand, though some HiFi DNA polymerases do not possess strand displacement activity. The same non-hairpin and long-hairpin designs were applied to Taq-based qPCR reactions. Due to the lack of 3’ to 5’ exonuclease activity, the quencher modifications on both companion strands were not digested by Taq DNA polymerase, regardless of the existence of the hairpin structure (Figure S3A and S3B). Moreover, the quencher strand on the long-hairpin companion strand functioned as a blocking modification to prevent Taq DNA polymerases from extending on the companion strand.

**Figure 1.**
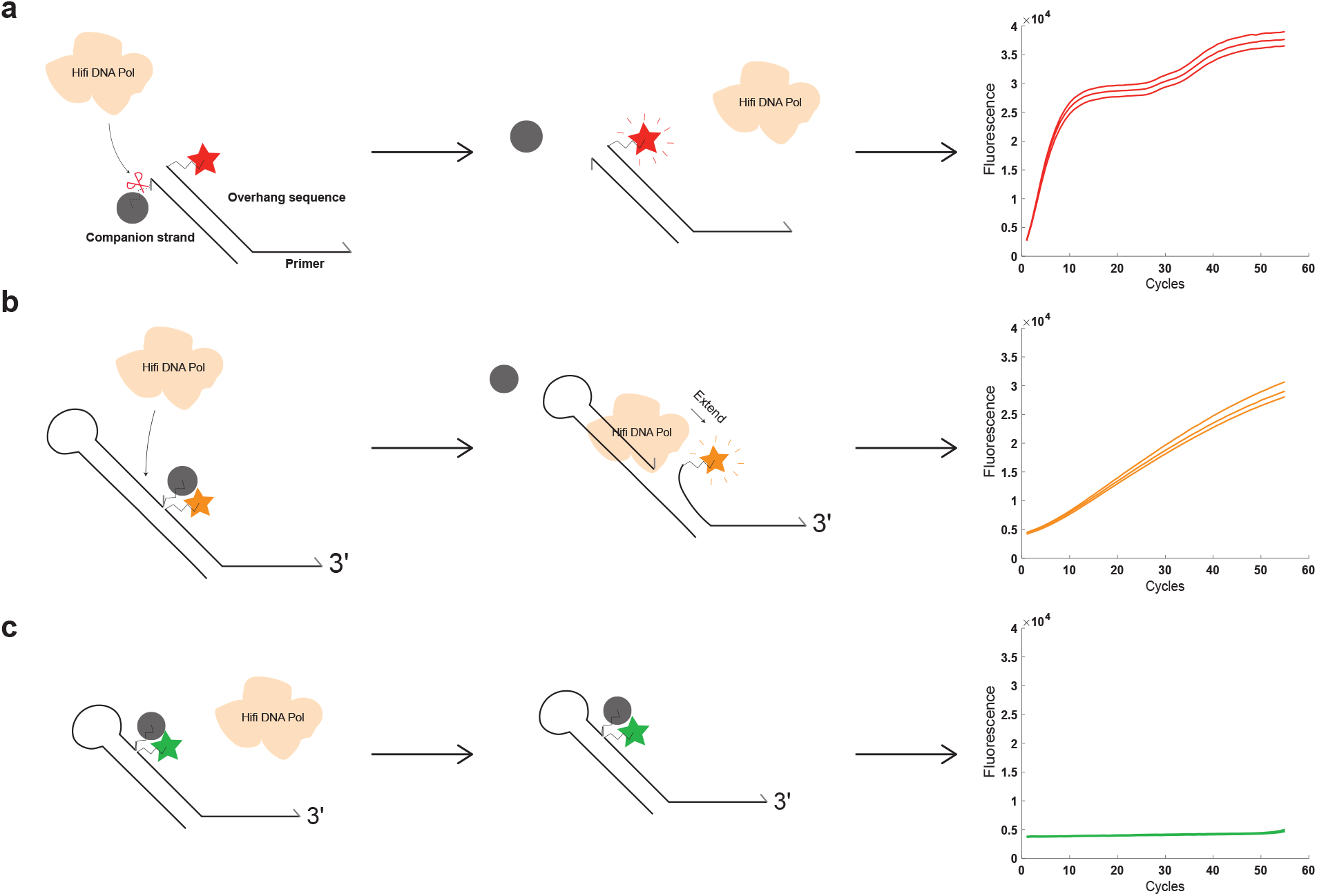
Hairpin structure effectively hindered the proofreading feature of high-fidelity (HiFi) DNA polymerases. **A). Chemical modification on the companion strand would be digested by HiFi DNA polymerases**. The companion strand is reverse complementary to the overhang sequence on the primer, and the 5’ overhang sequence is modified with a fluorophore modification. The companion strand has a 3’ quencher modification. This design failed to function as a fluorescence indicator, because the HiFi DNA polymerases would digest the 3’ quencher modification, resulting in abnormal increased signal even with the absence of template sequences. The untreated raw fluorescence signal detected by the CFX96 qPCR instrument was plotted in the right panel. The fluorescence signal that started above 0 as the background fluorescence indicated the slight unbalanced stoichiometric ratio of fluorophore probe and quencher probe, which would not influence the result interpretation. **B). Long-stem hairpin structure on the companion strand**. When applying a hairpin structure with 20nt stem to the 3’ of the companion strand, the long stem structure of the hairpin provided a binding site for the HiFi DNA polymerases, resulting in linearly increased fluorescent signal at the absence of template sequences, which may result from the digestion of quencher and the extension on the companion strand. The untreated raw fluorescence signal detected by the CFX96 qPCR instrument was plotted in the right panel. **C). Hairpin with a short stem reveals optimal performance for preventing self-extension and 3’ digestion**. Adjusted to a 4nt-stem structure, the hairpin on the 3’ of the companion strand promisingly functions as a fluorescence indicator. The 3’ to 5’ exonuclease activity of HiFi DNA polymerases on the quencher modifications was effectively prohibited. When there is no template present, there is not any abnormal fluorescence generated. The untreated raw fluorescence signal detected by the CFX96 qPCR instrument was plotted in the right panel.

By shortening the length of the stem to 4nt, we finally found a promising hairpin structure, which can sufficiently protect the 3’ region of the companion strand from the exonuclease digestion and polymerase extension within HiFi qPCR reactions (Figure 1C, Figure S3C). The phosphorothioate bonds (PTO) modification which was reported to resist proofreading activity^51^ and short-stem hairpin structure on the primer strand were analyzed and reported not compatible with HiFi qPCR reactions as fluorescence indicator (Figure S4).

### Overview of the Occlusion System

The Occlusion System is composed of an Occlusion Primer and an Occlusion Probe, the Occlusion Probe is originated from the companion strand described above (Figure 2A). As the HiFi DNA polymerase extended the reverse primer on the strand newly synthesized from the Occlusion Primer, the separation of fluorophore and quencher led to the production of fluorescent signals. We demonstrated the feasibility of the Occlusion System in detecting the single target by adding the template genomic DNA (Figure 2B). Furthermore, we interchanged the quencher and fluorophore modifications and re-applied the design in qPCR reactions (Figure 2C). We continued to explore the multiplexing ability of the Occlusion System in qPCR reactions. By randomly mixing each two or three sets of Occlusion oligos together, we demonstrated the combability of the Occlusion System functioning as the fluorescence indicator in multiplex HiFi qPCR reactions (Table 1). The closeness of the Ct value under single-plex or multi-plex revealed that the multiplexed Occlusion System sets could indicate the generation of multiple amplicons without influencing the PCR efficiency.

**Figure 2.**
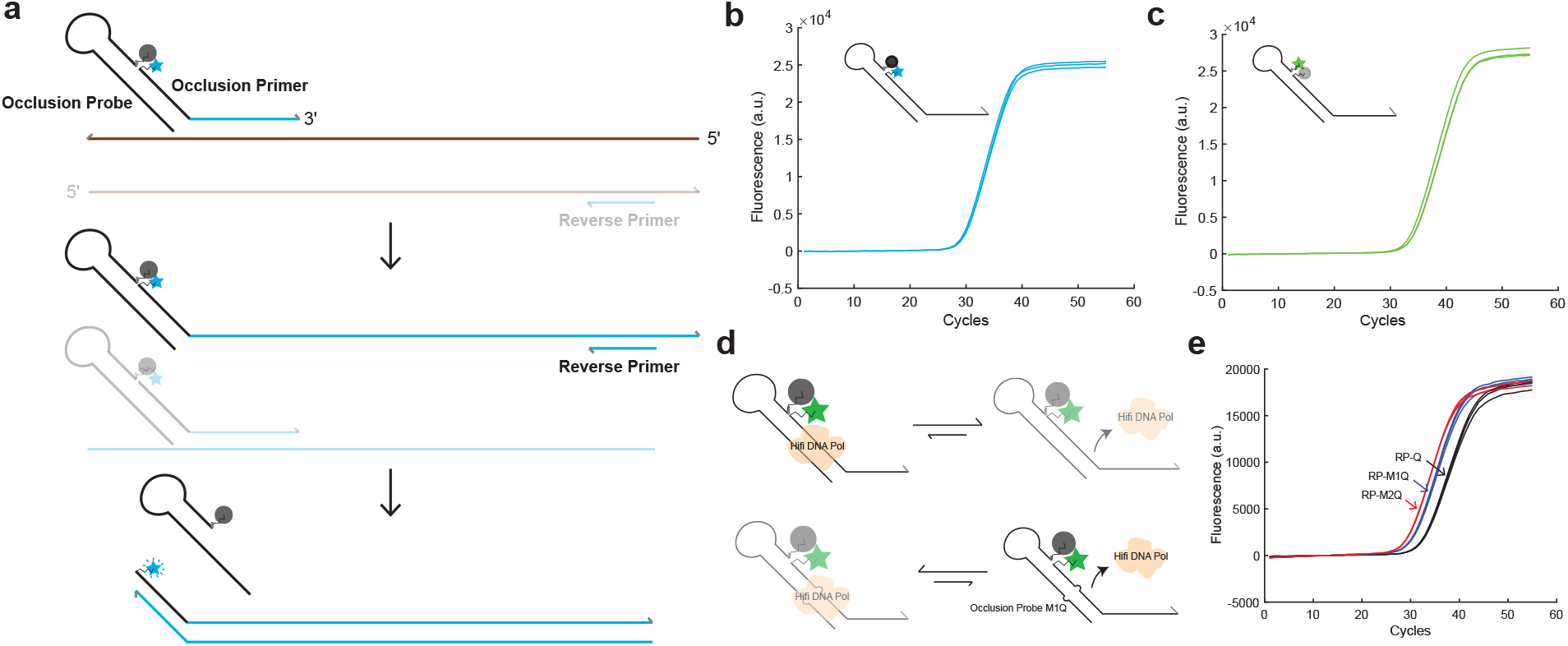
Overview of the Occlusion System. **A). Schematic of the Occlusion System**. The Occlusion System mainly comprises an Occlusion Probe and an Occlusion Primer. The Occlusion Probe is originated from the companion strand described in Figure **1C**. The Occlusion Primer has a primer region and a 5’ overhang region modified with fluorophore. The Occlusion Primer could bind to the template sequence and extend on it. As the Reverse Primer extends on the newly synthesized strand from the Occlusion Primer, it would encounter and then displace the Occlusion Probe, causing the separation of the fluorophore and quencher, thus, the fluorescent signal would increase, indicating the generation of amplicons. **B). Fluorescence signal of a typical Occlusion single-plex qPCR reaction using HiFi polymerase**. Usually, there would be a fluorophore modification on the Occlusion Primer, a quencher modification on the Occlusion Probe. **C). Design of switched fluorophore and quencher modification**. The switch of fluorophore and quencher modifications would not affect performance of the Occlusion system. **D). Schematic of mismatches in releasing occupied DNA polymerases**. The double-stranded region between the Occlusion Primer and the Occlusion Probe provides binding site for the DNA polymerases, resulting in the occupancy of DNA polymerases and the reduced amount of effective DNA polymerases in PCR amplification. **E). Comparison of qPCR curves between Occlusion probes with different mismatches**. By comparing the qPCR results from Occlusion Probe without mismatch, the Occlusion Probe with single mismatch, and the Occlusion Probe with double mismatches, we concluded that the introduction of the mismatches on the Occlusion system improve the PCR efficiency more. Also, the increase of the total number of the mismatches benefits the overall PCR efficiency.

**Table 1.** Ct summary of multiplexed Occlusion qPCR reactions. By designing three sets of Occlusion Primer and Occlusion Probe to target three genes (*FLT3, IDH1* and *DNMT3A*), the ability of the Occlusion System to indicate amplification of multiplex targets within the same reaction was demonstrated.

| Information of the input | Ct of Cy5 | Ct of HEX | Ct of ROX |
| --- | --- | --- | --- |
| **Cy5** | 24.9 | Inf | Inf |
| **HEX** | Inf | 25.2 | Inf |
| **ROX** | Inf | Inf | 26.2 |
| **Cy5 + ROX** | 25.2 | Inf | 27.2 |
| **Cy5 + HEX** | 25.1 | 25.4 | Inf |
| **HEX + ROX** | Inf | 25.5 | 26.6 |
| Cy5 + HEX + ROX | 25.3 | 25.7 | 27.6 |

Moreover, we hypothesized that the double-stranded region formed between the overhang of the Occlusion Primer and the Occlusion Probe would provide extra binding sites to the DNA polymerases, resulting in unnecessary polymerase occupancy and reduced PCR efficiency. Mismatch(es) was introduced to every 10 to 20 nucleotides^49^ within the double-stranded region to solve this problem (Figure 2D). As shown in Figure 2E, the dual-nucleotide-mismatched Occlusion Probe revealed the minimal Ct value, showing the PCR efficiency was improved by reducing the unnecessary polymerase occupancy.

Then we applied the Occlusion System to the BDA technology, as the schematic was shown in Figure S5. The Occlusion System is compatible with diagnostic methods like the BDA using HiFi DNA polymerases in the absence of any other fluorescence indicator. The result revealed the non-overlapping fluorescence signal among samples with different VAFs and clear background with imperceptible noise in the fluorescent channel (Figure 3A, Figure S6). Due to the closeness between 0.01% VAF and the wildtype genomic DNA for the two *IDH1* mutations and the *FLT3* 2503 G>T mutation, further statistical analysis was conducted on 24 repeats on the 0.01% VAF and wildtype genomic DNA of these mutations (Figure S7 and S8). Compared with the reactions using Taq DNA polymerase, each BDA design reveals 3- to 10-fold improvement in LoD when using HiFi DNA polymerases (Figure S9). To see if the Occlusion System allows multiplexed BDA readout in qPCR reactions, we designed different BDA Occlusion sets for each of the two target genes of interest (Figure 3B). Both channels for each pair could achieve effective variant enrichment. Each 10% and 1% VAF sample was NGS quantified (Table S1), and each 0.01% VAF sample was Sanger-sequenced post reaction (Figure S11 and S12). In this way, we showed the Occlusion System is capable of multiplexing qPCR reactions, and compatible with diagnostic methods like BDA, achieving ultra-sensitive variant enrichment with VAF down to 0.01% in HiFi qPCR.

**Figure 3.**
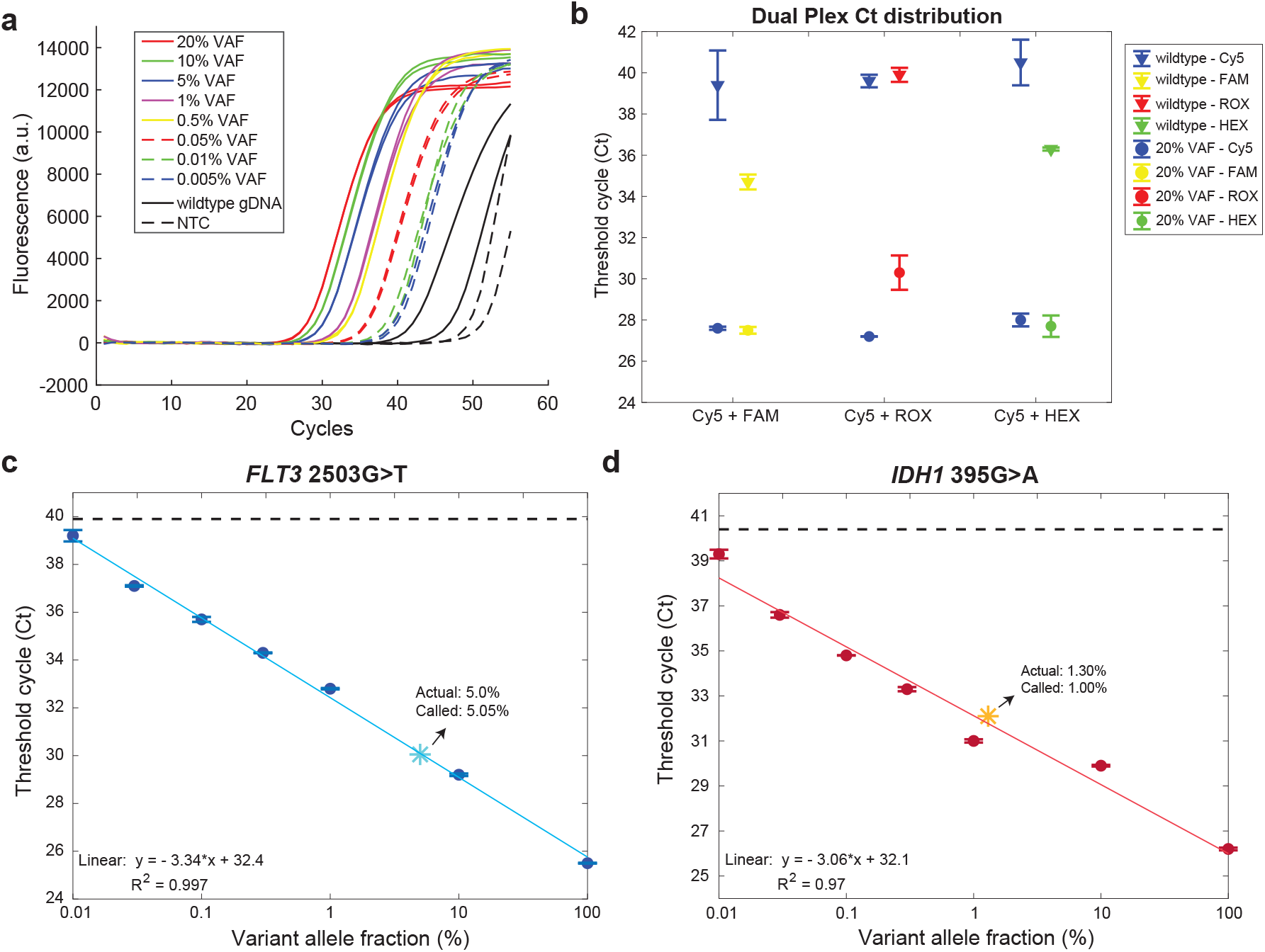
The Occlusion System with blocker displacement amplification (BDA) technology. **A). The single-plex BDA Occlusion reaction**. The compatibility of the Occlusion System with the BDA technology by applying the Occlusion System to the reverse primer of the BDA design, as shown in Figure **S5**. Duplicate qPCR results for various mixtures of synthetic gBlock variants and genomic DNA with different VAFs. Input 200 ng or equivalent to 60,000 copies of molecules per reaction. **B). The multiplexed BDA Occlusion reaction**. By combining two BDA Occlusion sets together inside one qPCR reaction, we demonstrated the multiplexing ability of the Occlusion System with BDA. The genes of interest here are *FLT3* and *DNMT3A*. Amplification of *FLT3* region was indicated by Cy5 channel. The FAM, ROX, and HEX channels stand for amplicons from *DNMT3A* gene. The variant templates used are the mixture of 20% VAF FLT3 2503G>C and 20% VAF *DNMT3A* 2644C>T. **C). Quantitation validation from standard VAF curve of the *FLT3* 2503G>T mutation with a 5.0% reference sample**. Ct value of each VAF sample was plotted as each individual blue dots with one standard deviation as error bar. The solid blue line was the fitted line, showing a strong linear correlation between log (VAF) and Ct values with an r-square of 0.997. The 5.0% VAF reference sample was marked as a light blue star, its coordinate was determined by the claimed VAF and the obtained Ct value from the Occlusion qPCR reaction. **D). Quantitation validation from standard VAF curve of the *IDH1* 395G>A mutation with a 1.30% reference sample**. The VAF samples were presented as individual red dots, with the fitted line shown as solid red. The linear correlation in *IDH1* 395G>A mutation has an r-square around 0.97. The 1.30% VAF reference sample was marked as a yellow star, its closeness to the fitted line revealed the accuracy of VAF calling.

### Clinical validation of the HiFi BDA Occlusion System

The analytical performance of Occlusion BDA systems was further explored. We plotted Ct values of each VAF sample as each data point versus logarithmic VAF values, to generate a fitted curve from linear regression. The fitted curves exhibited a strong linear correlation with r-squares ranging from 0.896 to 0.999 (Figure 3C and 3D, Figure S10). To further validate whether the fitted curve generated from the mimicking VAF samples is appliable and accurate enough for calculating VAF from clinical specimens, we designed BDA Occlusion sets for *IDH1, FLT3, DNMT3A* gene targets and ran commercial reference samples on these genes. The closeness of the reference samples (shown as the light blue star in Figure 3C, orange star in 3D, and magenta star in S10A) to the fitting curve exhibits the accuracy of the fitting result. The percentage error between the called VAF and claimed VAF is 1% for 5% reference sample, 23.1% and 20.8% for 1.30% reference sample. We expect the sample with lower VAF would have more randomness with the variation from the number of input variant sequences based on Poisson distribution.

To demonstrate the compatibility with clinical samples, we performed HiFi BDA Occlusion qPCR reactions on the 200 ng genomic DNA extracted from PBMC samples of AML patients and healthy individuals. We used the *GAPDH* housekeeping gene to quantify the input of samples, a summary of Ct value was shown in Table S4 to S5. Then we applied *FLT3* and *IDH1* BDA Occlusion designs in the DNA samples extracted from all AML patients and healthy donors, with 6 *FLT3* mutations, 2 *IDH1* mutations reported from 7 individual AML samples (Figure S13, detailed Ct values were collected and summarized in Table S6 to S9). By confirming the mutation pattern through Sanger sequencing (Figure 4B and S14), we successfully called the VAFs of these samples based on the Ct value and the fitted VAF curve of the corresponding mutation (Table S10). Considering the limited sample resource in clinical diagnosis, we further downscaled the input to roughly 20 ng to 30 ng per *FLT3*-positive sample on the HiFi BDA Occlusion qPCR and the ddPCR platform (Figure 4A, Figures S15 to S19, Table S11). As shown in Figure 4A, there was 100% concordance between the VAFs called by the ddPCR and the HiFi BDA Occlusion System in the *FLT3* mutations with VAF higher than the corresponding LoDs. Thus, we concluded that the HiFi BDA Occlusion System was accurate and sensitive in identifying VAF from unknown clinical samples, showing advantages in detecting mutations down to 0.01% VAF with flexible and scalable DNA input, and the results were fully comparable with gold standard ddPCR, indicating a promising application in facilitating clinical diagnosis with a fast turnaround time.

**Figure 4.**
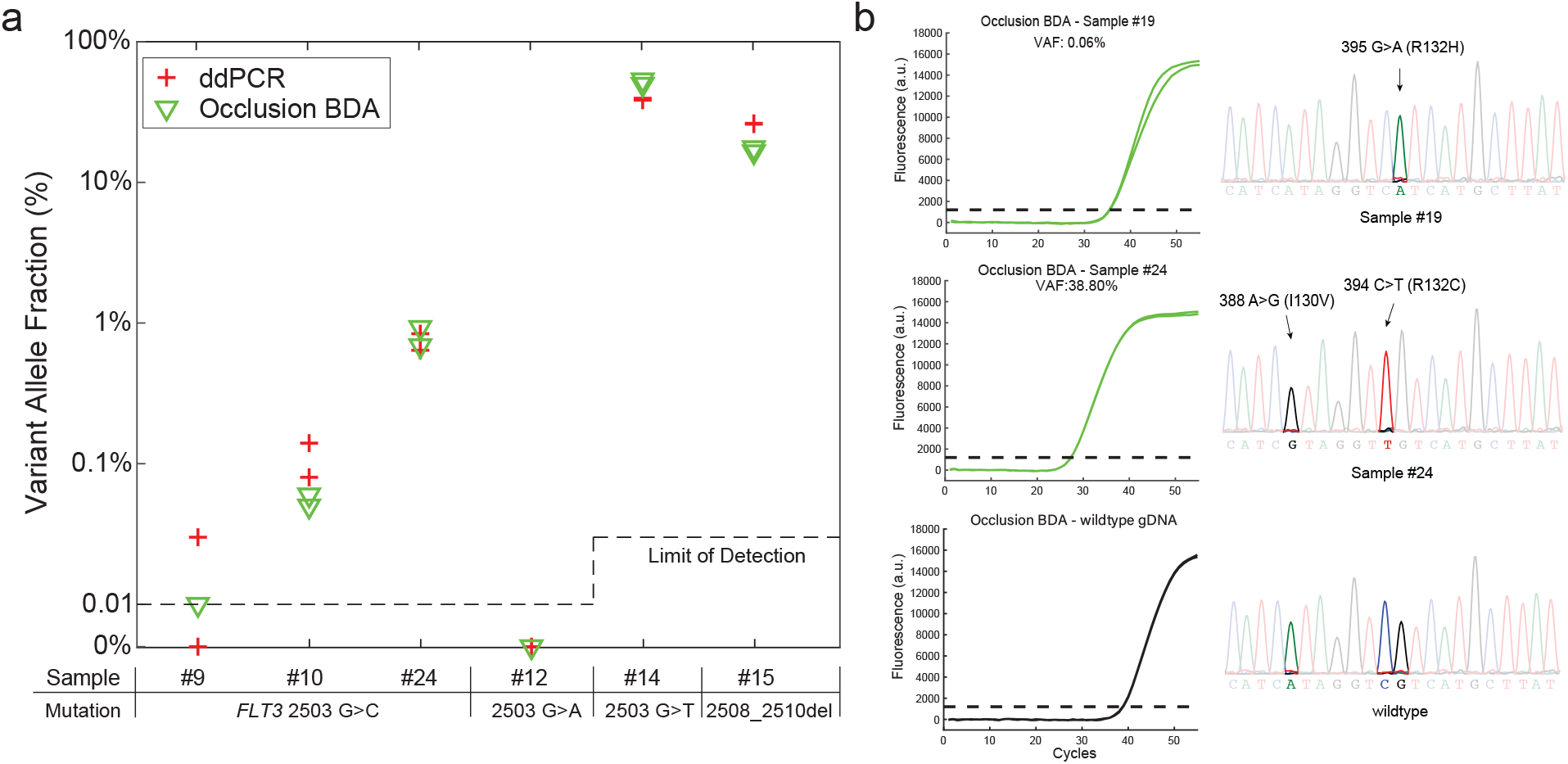
Ct determination validation with commercial clinical samples. **A). Summary of comparative results between ddPCR and HiFi BDA Occlusion qPCR in *FLT3* mutations**. Each sample was duplicated in both ddPCR and qPCR reactions. The results showed 100% concordance between ddPCR and qPCR reactions. Note that, for sample #9, one of the dupplicate ddPCR reactions failed to report mutation. **B). HiFi BDA Occlusion qPCR results and Sanger traces of AML samples reported positive in the *IDH1* gene**.

## DISCUSSION

This Occlusion System presented here, with rationally designed short-stem hairpin structures, could hinder the 3’ to 5’ exonuclease activity of HiFi DNA polymerases on the 3’ chemical modification (the fluorophore or quencher modification), and function as the probe-based fluorescence indicator in single- and multi-plex HiFi qPCR reactions. Although the structure and feature of HiFi DNA polymerases have been extensively researched^30,35–38,52–54^, we are not aware of any other probe-based designs reported to be compatible with HiFi DNA polymerases as the fluorescence indicator in multiplex qPCR reactions. The high polymerase misincorporation rate from the lack of proofreading feature of the non-HiFi DNA polymerases has been a restriction factor to the detection sensitivity of many common variant detection methods. It is urged to upgrade current variant detection methods or develop new methods which are easy-to-operate and low-in-complexity to satisfy the clinical requirements for better disease diagnosis and monitoring. The appearance of HiFi DNA polymerases, with the fidelity was improved by 50- to 250-fold through the proofreading activity, offered a potential solution by being easily adapted to the current variant detection methods. The Occlusion System in this work provided a simple and easy-to-apply probe-based design to efficiently indicate the generation of amplicons in a flexible combination of numbers of fluorescence channels, which overcomes the current bottleneck for extensively applying HiFi DNA polymerases in the multiplex real-time reactions.

This Occlusion System demonstrated its compatibility with multiplexed variant enrichment BDA reactions with HiFi DNA polymerases. It allows the convenient transition from previously non-high-fidelity DNA polymerases to high-fidelity DNA polymerases for other qPCR-based variant detection technologies. This convenience translates into practical advantages for molecular diagnostics and genomics research, allowing previous variant detection methods to achieve better sensitivity and specificity, and enabling detection of multiple targets within the same reaction. We showed that the Occlusion system with the BDA technology could detect multiplexed variants with VAF low to 0.01%, which is up to 10-fold improvement compared with the LoD using non-HiFi DNA polymerases. The BDA technology has been mostly reported to have an LoD of only around 0.1% VAF, except As-BDA which could detect mutation low to 0.01% VAF. However, As-BDA requires preliminary knowledge of mutation or small insertion/deletion information, thus limited in the total number of mutations detected within one reaction and restricted in discovering novel mutations. The HiFi BDA Occlusion system offsets this limitation by achieving a hypothesis-free variant enrichment within the target region with ultra-sensitivity, and the specific mutation identity could be confirmed by Sanger sequencing or other sequencing technologies. Furthermore, the same sequence of Occlusion probe could be re-used in designs targeting different templates within the same fluorescent channel, reducing the overall cost in oligo synthesis. Thus, we believe the Occlusion System provides a cost-efficient, user-friendly, and plex-scalable solution in high-fidelity DNA qPCR reactions, which is not achievable by other methods.

We expect the Occlusion system can promote the diagnosis of early-stage cancer clinically. In this work, we primarily designed the HiFi BDA Occlusion System to detect mutations that occurred within codon 126 to codon 133 in the *IDH1* gene and codon 832 to codon 838 in the *FLT3* gene. By applying the HiFi BDA Occlusion qPCR reactions in PBMC samples from 27 AML patients and 15 healthy donors, we successfully detected mutations in 7 AML samples and confirmed the results by Sanger sequencing. When compared with ddPCR, results are 100% concordant for mutations with VAF ≥ 0.1% with the similar DNA input. When it comes to lower VAF mutations, unlike ddPCR which is less dynamic in the input with the concern of multiple molecules within the same droplet, the HiFi BDA Occlusion system successfully detected two samples with VAF below 0.05% using 10-fold more DNA input. The HiFi BDA Occlusion system has demonstrated its reliable capability, robustness and competitiveness in detecting VAF low to 0.01% in clinical PBMC samples, which reveals the potential of widespread applications in many other clinical specimen types. We also believed that the Occlusion system was a general method that could be used with many other variant detection methods in high-fidelity PCR reactions.

Besides the application in HiFi qPCR reactions, we are confident that the Occlusion System can replace TaqMan probes or other probe-based fluorescence indicators in non-HiFi qPCR reactions. There are limitations in designing and applying TaqMan probes within competitive genomic regions or ultra-short amplicons, the Occlusion System is more suitable as the fluorescence indicator in these scenarios.

## Data Availability

The sequences of the DNA oligos used for experiments are included in the supplementary file accompanying this manuscript. The detailed concentration of the oligos used for experiment are described in the Method and Materials section. The main data supporting the results in this study are available within the paper and its Supplementary Information. The datasets collected and/or analyzed during the current study available from the corresponding author on reasonable request.

## ACKNOWLEDGEMENTS

Correspondence may be addressed to DYZ.

The authors thank Quoc-Khanh Pham and Dr. Bao for use of their ddPCR instrument.

The authors thank Xuwen Li, Gavin Jiaming Li and Nina Guanyi Xie for editorial assistance.

## AUTHOR CONTRIBUTIONS

K.Z and D.Y.Z conceived the project. K.Z, A.P, L.R and C.W performed the experiments. K.Z, A.P, M.W analyzed the data. K.Z, A.P, L.Y.C, P.D and P.S conducted Occlusion design. K.Z wrote the manuscript with input from all authors. D.Y.Z. revised the manuscript.

