## Supplementary file for "Hairpin structure facilitates multiplex high-fidelity DNA amplification in real-time PCR"

**Section S1. Impact of different DNA polymerases on the limit of detection (LoD) for variant detection method. 2**

**Section S2. Performance of high-fidelity DNA polymerases with previous probe designs. 3**

**Section S3. Further exploration on the Occlusion System. 4**

**Section S4. The Occlusion System with BDA. 6**

**Section S5. The Occlusion qPCR and ddPCR results of PBMC samples from healthy donors and AML patients. 13**

**Section S6. List of the Occlusion Probe component concentration and oligonucleotide sequences. 24**

**Section S1. Impact of different DNA polymerases on the limit of detection (LoD) for variant detection method.**

The limit of detection (LoD) of the same Blocker displacement amplification (BDA)[1] design could be different with distinct DNA polymerases used, as shown in Figure S1. The same BDA design has been applied to the single-plex qPCR reactions using Taq DNA polymerase (Figure S1A) or Phusion DNA polymerase (high-fidelity DNA polymerase, Figure S1B). The targeted mutation here is *DNMT3A* R882C (2644C>T).

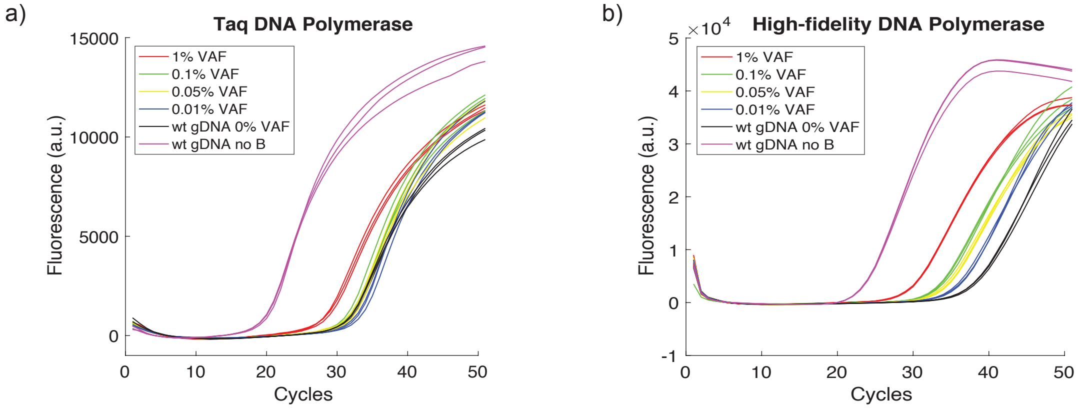


**Figure S1. Limit of Detection with different DNA polymerase.**

**(A). LoD with Taq DNA polymerase.** The sensitivity of BDA reactions using Taq DNA polymerases would be limited in 1% VAF. Each reaction of the triplicate has around 200ng DNA input. The Taq DNA polymerase used here is PowerUp SYBR Green Master Mix.

**(B). Lod with high-fidelity DNA polymerase (Phusion).** The sensitivity of BDA reactions using Phusion high-fidelity DNA polymerases would be down to 0.01% VAF. Each reaction of the triplicate has around 200ng DNA input. SYTO 13 green-fluorescent nucleic acid stain is used to indicate the generation of amplicons.

**Section S2. Performance of high-fidelity DNA polymerases with previous probe designs.**

To see if previous probe designs, which work well with Taq DNA polymerases, were able to work with high-fidelity DNA polymerases, we applied Phusion DNA polymerase in reactions with TaqMan probes and Sunrise primer respectively (Figure S2).

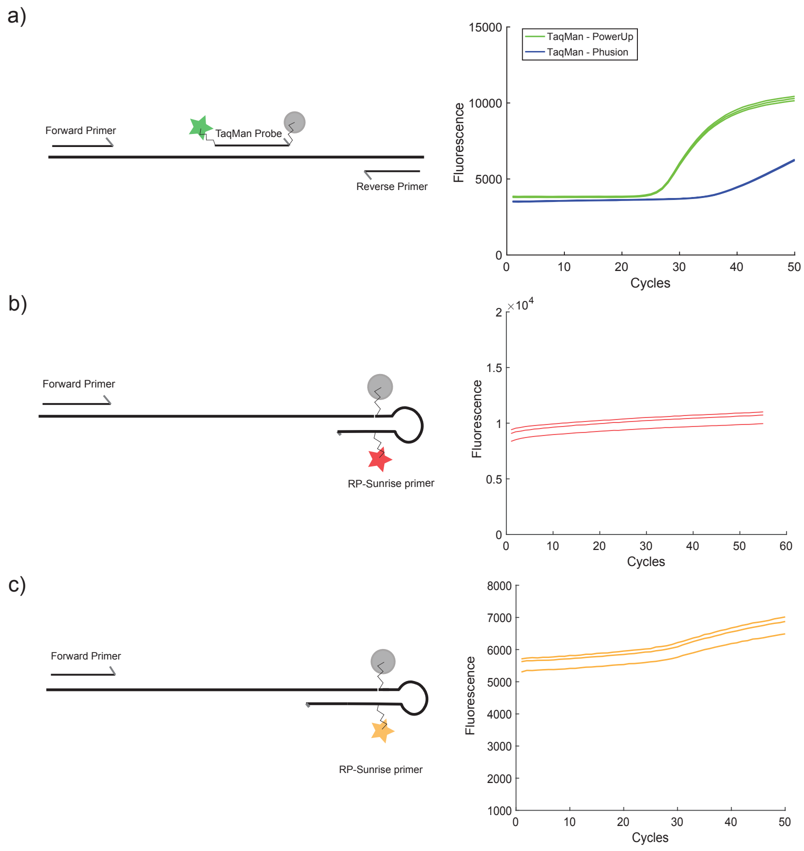


**Figure S2. Non-compatibility of TaqMan probes and Sunrise primers with high fidelity DNA polymerases.**

**(A). High-fidelity DNA polymerases with TaqMan probe.** Compared with the fluorescence post-subtraction from the background signal and Ct value in reactions with PowerUp DNA polymerase (Taq), the fluorescence in reactions using Phusion DNA polymerases was much lower, and the overall Ct value was delayed for approximately 10 cycles. Thus, TaqMan lost its sensitivity when combined with high fidelity DNA polymerases. Each reaction was performed in triplicate.

**(B) and (C). High-fidelity DNA polymerases with Sunrise probe.** The raw fluorescent signal presented showed little or no amplification curve in high-fidelity DNA polymerases, indicating the incapability of Sunrise primers with high-fidelity DNA polymerases. Each reaction was performed in triplicate.

**Section S3. Further exploration on the Occlusion System.**

When applying the same set of designs as shown in the Figure **1** in qPCR reactions using Taq DNA polymerase which does not possess 3' to 5' exonuclease activity, there was no abnormal fluorescence signal generated, indicating the generation of abnormal signal in HiFi qPCR reactions was from the proofreading activity of the HiFi DNA polymerases (Figure S3). More designs on the companion strand were evaluated and shown in Figure S4.

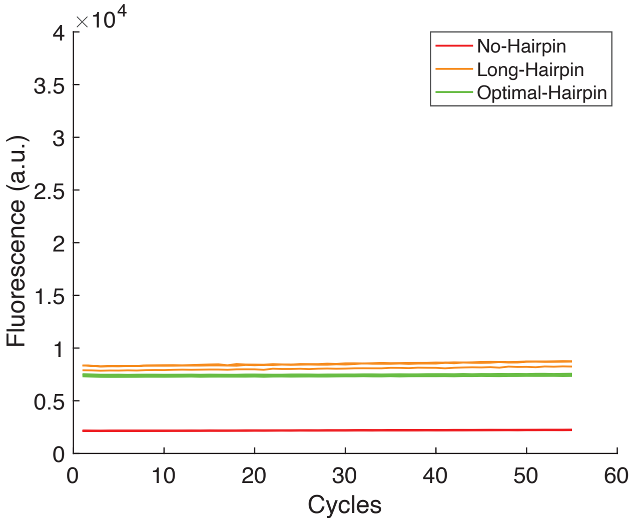


**Figure S3. Performance of different designs of companion strands with Taq DNA polymerase.**

**A-C). qPCR curves of each reaction with different design input.** The same designs shown in the Figure **1** were input in Taq qPCR reactions. Due to the lack of 3' to 5' exonuclease, the Taq DNA polymerase did not produce any abnormal fluorescence signal at the absence of template. The untreated raw fluorescence signal detected by the CFX96 qPCR instrument was plotted in the right panel. The fluorescence signal that started above 0 as the background fluorescence indicated the slight unbalanced stoichiometric ratio of fluorophore probe and quencher probe, which would not influence the result interpretation.

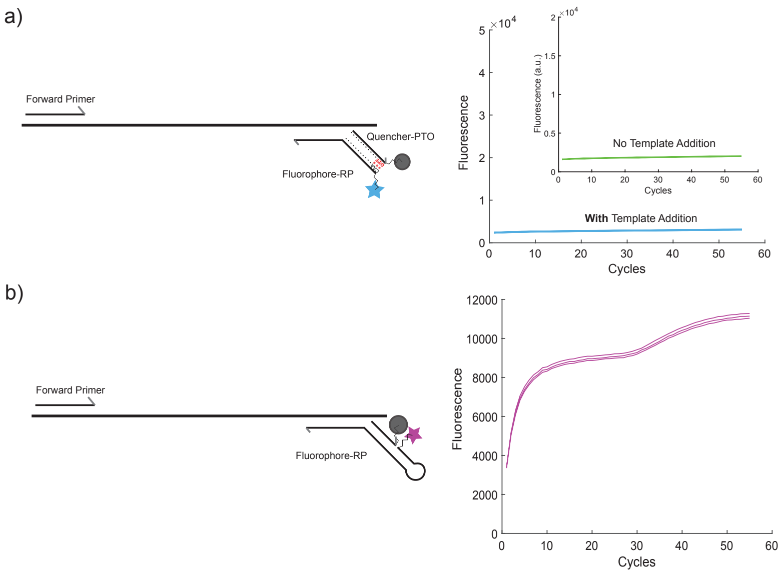


**Figure S4. Experimental validation of other designs to the companion strand with the addition of template.**

**(A). Phosphorothioate bonds modifications (PTO) design.** When applying the PTO modification on the companion strand, the PTO modifications inhibited the proofreading feature on the quencher modification and avoided the abnormal fluorescence signal. However, the reaction failed to generate any fluorescence signal even with the template added, indicating the non-compatibility of PTO modification with the companion strand.

**(B). Hairpin structure on the companion strand.** When applying the hairpin structure on the primer strand rather than the companion strand, the hairpin structure lost its prohibition ability in the proofreading feature. This demonstrated the necessity of putting hairpin structure in the 3' of the companion strands.

**Section S4. The Occlusion System with BDA.**

To further demonstrate if the Occlusion System is compatible with variant detection methods, we applied Occlusion system with BDA[1] technology. The detailed schematic is presented in Figure S5.

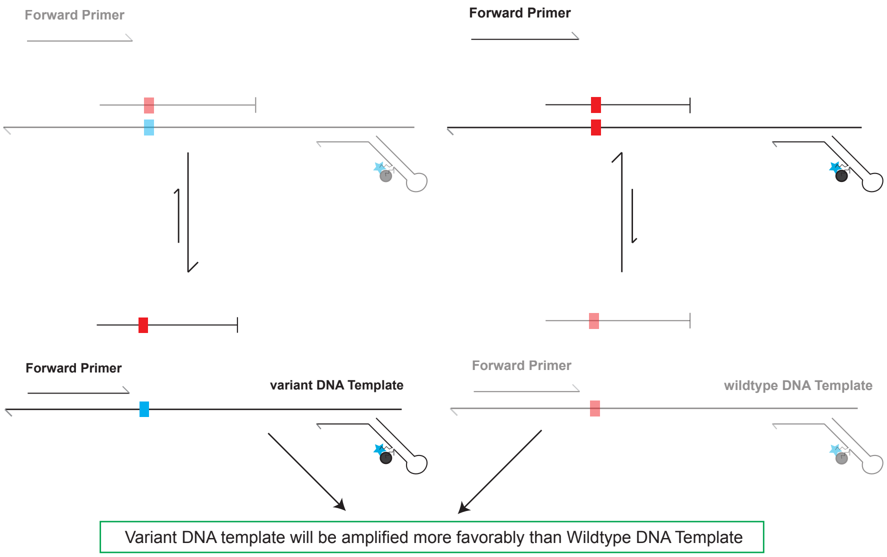


**Figure S5. Schematic of BDA Occlusion System.** In the BDA Occlusion reactions using high fidelity DNA polymerases, the normal reverse primer was replaced with the Occlusion Primer. Additionally, An Occlusion Probe would be used to pair with the Occlusion Primer. Then they would be mixed with BDA forward primer and BDA blocker which are the same as in a typical BDA reaction. In qPCR reactions, the Occlusion Probe and the Occlusion Primer could be chemically modified with fluorophores and quenchers respectively to indicate amplicon generation.

We then applied the BDA Occlusion designs in multiple genes for qPCR reactions with both Taq DNA polymerases and high-fidelity DNA polymerases, as shown in Figure S6 to S9. The separation in Ct value of each VAF sample proved the ability of 0.01% VAF detection. Statistical analysis on Ct values of 0.01% VAF samples and wildtype samples were shown in Figure S7 and S8, with each sample repeated for 24 reactions (the wildtype sample shown in Figure S7 were repeated for 47 reactions). Then Ct values of each duplicate reaction of each synthetic template were plotted versus logarithmic VAF values, and liner correlation was calculated based on curve fitting (Figure S10). The reaction products were sent to Sanger sequencing for further sequence validation (Figure S11 and S12).

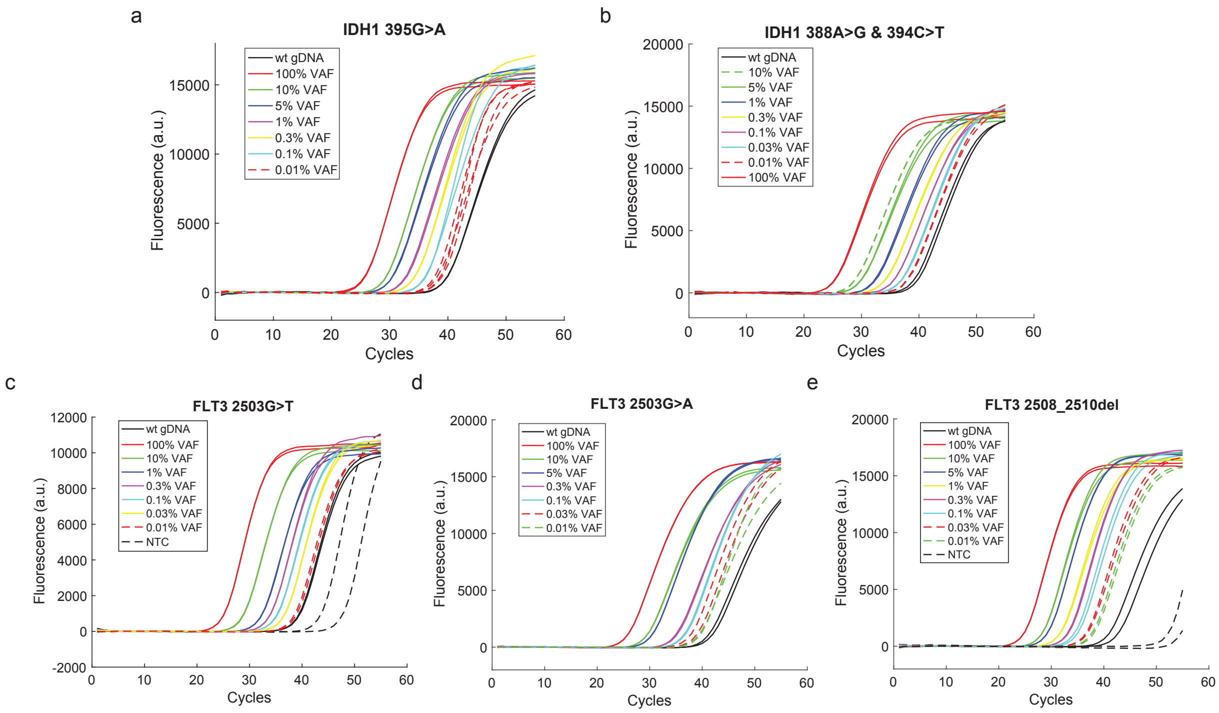


**Figure S6. qPCR curves for each BDA Occlusion design using high-fidelity DNA polymerases.**

**(A). qPCR result for *IDH1* 395G>A mutation.** Various mixtures of synthetic gBlock variants and genomic DNA with different VAFs were input, each mixture was performed in duplicate except the 0.01% VAF sample were performed in quadruplicates. Further statistical analysis on the Ct values of 0.01% VAF sample and the wildtype sample were shown in Figure S7.

**(B). qPCR result for *IDH1* 388G>A (I130V) & 394C>T mutation.** Each mixture was performed in duplicate. Further statistical analysis on the Ct values of 0.01% VAF sample and the wildtype sample were shown in Figure S7.

**(C). qPCR result for *FLT3* 2503G>T mutation.** Each mixture was performed in duplicate. Further statistical analysis on the Ct values of 0.01% VAF sample and the wildtype sample were shown in Figure S8.

**(D). qPCR result for *FLT3* 2503G>A mutation.** Each mixture was performed in duplicate.

**(E). qPCR result for *FLT3* 2508_2510del (I836del) mutation.** Each mixture was performed in duplicate.

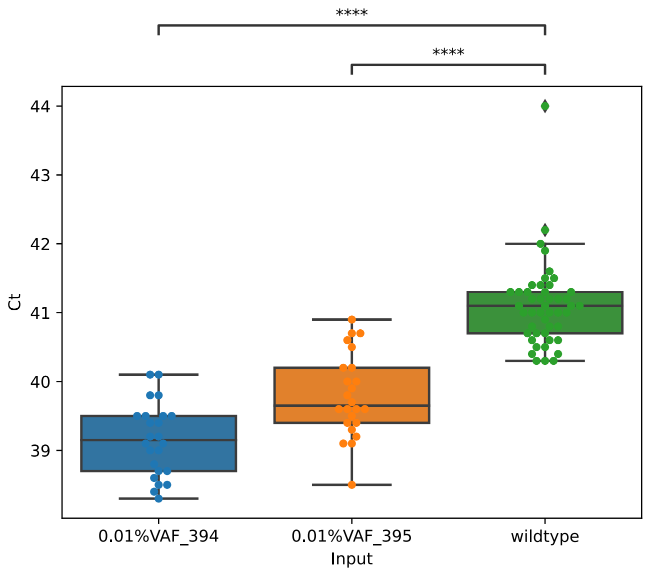


**Figure S7. Statistical analysis on Ct values of 0.01% VAF samples and the wildtype sample for two mutations in the *IDH1* gene.** Ct values of 24 reactions for each input (47 reactions for the wildtype) were collected and the p-value was analyzed using independent t-test.

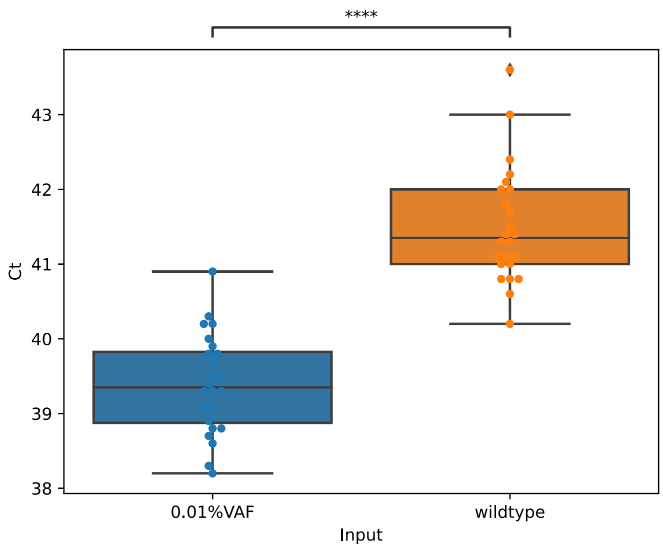


**Figure S8. Statistical analysis on Ct values of 0.01% VAF samples and the wildtype sample for 2503 G>T mutation in the *FLT3* gene.** Ct values of 24 reactions for each input were collected and the p-value was analyzed using independent t-test.

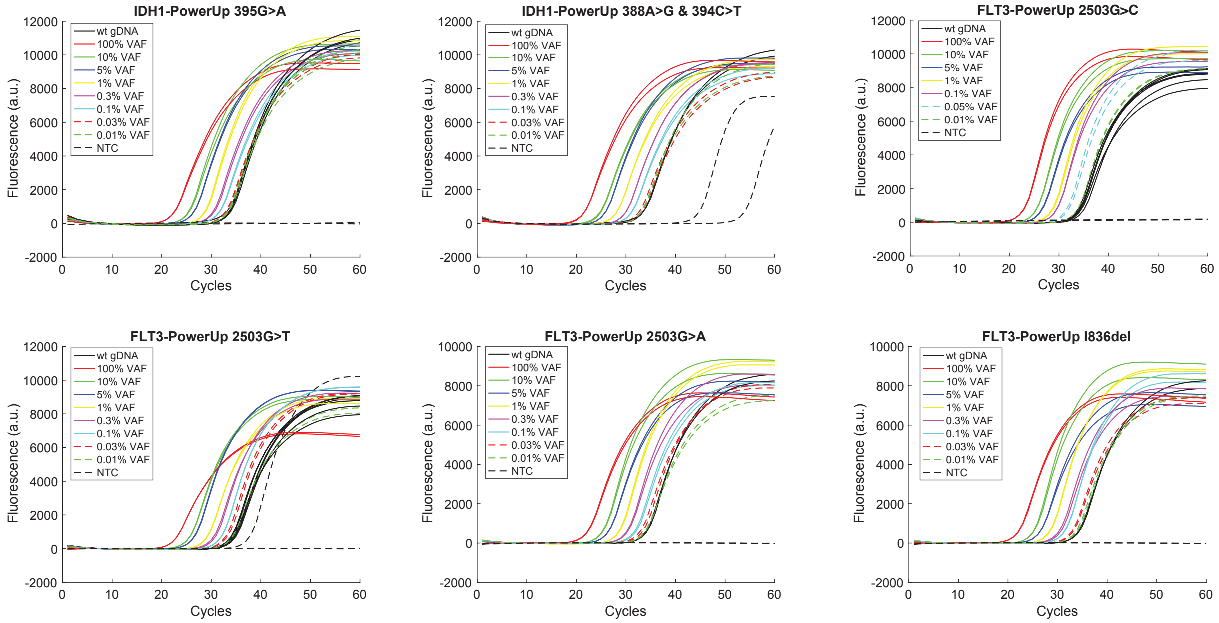


**Figure S9. qPCR curves for each BDA Occlusion design using Taq DNA polymerases.**

**(A). qPCR result for *IDH1* 395G>A mutation.** There reveals no difference for the Ct values among 0.03% VAF and the wildtype sample, demonstrating the LoD is restricted to 0.1% VAF, which is 10-fold less than that using HiFi DNA polymerases.

**(B). qPCR result for *IDH1* 388G>A (I130V) & 394C>T mutation.** There reveals no significant difference for the Ct values among 0.03% VAF and the wildtype sample, demonstrating the LoD is 0.1% VAF, which is 10-fold less than that using HiFi DNA polymerases.

**(C). qPCR result for *FLT3* 2503G>C mutation.** There reveals no significant difference for the Ct values among 0.01% VAF and the wildtype sample, demonstrating the LoD is 0.05% VAF, which is 5-fold less than that using HiFi DNA polymerases.

**(D). qPCR result for *FLT3* 2503G>T mutation.** There reveals no significant difference for the Ct values among 0.01% VAF and the wildtype sample, demonstrating the LoD is 0.03% VAF, which is 3-fold less than that using HiFi DNA polymerases.

**(E). qPCR result for *FLT3* 2503G>A mutation.** There reveals no significant difference for the Ct values among 0.01% VAF and the wildtype sample, demonstrating the LoD is 0.03% VAF, which is 3-fold less than that using HiFi DNA polymerases.

**(F). qPCR result for *FLT3* 2508_2510del (I836del) mutation.** There reveals no significant difference for the Ct values among 0.01% VAF and the wildtype sample, demonstrating the LoD is 0.03% VAF, which is 3-fold less than that using HiFi DNA polymerases.

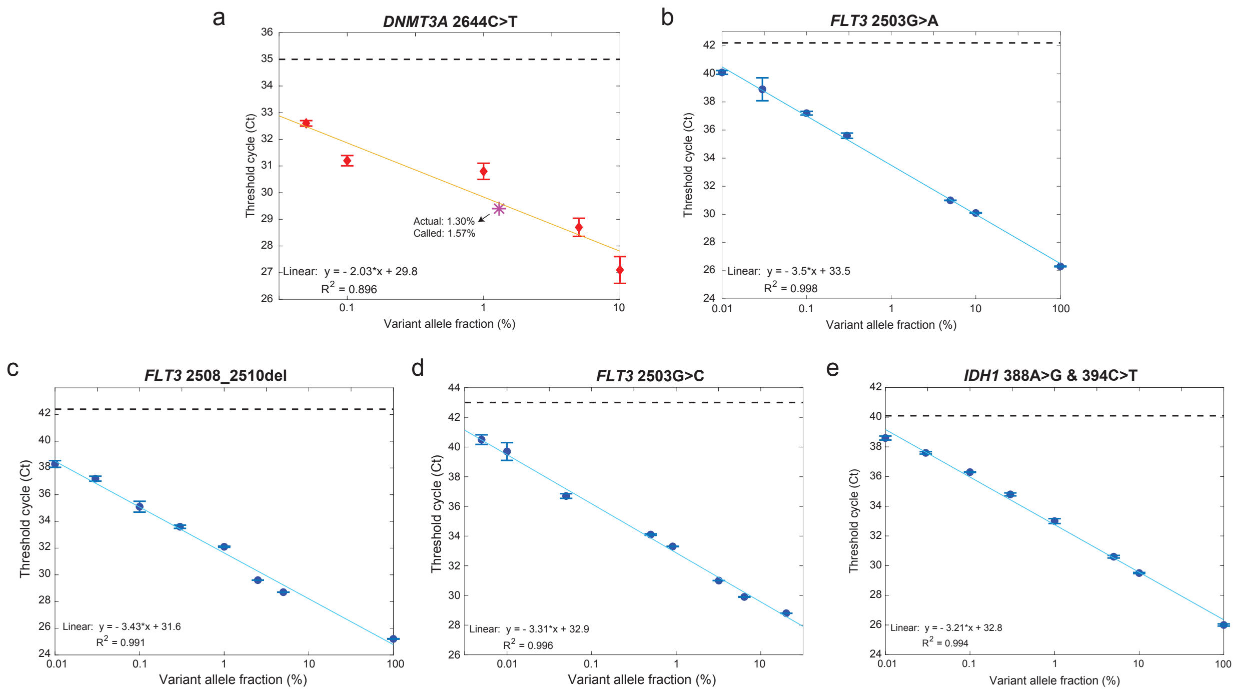


**Figure S10. Standard VAF curve of mutations.**

**(A). Standard VAF curve of 2644C>T (R882C) mutation in *DNMT3A* gene.** Each data point was from a duplicate reaction and presented as individual red dot. The error bar stands for one standard deviation. The fitted line is yellow. The 1.30% VAF reference sample was marked as a magenta star, whose coordinates was determined by its Ct value and the vendor claimed VAF. The closeness of the reference sample to the fitted line exhibits the accuracy of the fitting result. The Ct values and log VAFs exhibited liner correlation with an r-square is 0.896.

**(B). Standard VAF curve of 2503G>A (D835N) mutation in *FLT3* gene.** Each data point was from a duplicate reaction and presented as individual blue dot. The error bar stands for one standard deviation. The fitted line is light blue. The Ct values and log VAFs exhibited liner correlation with an r-square is 0.991.

**(C). Standard VAF curve of 2508_2510del (D835H) mutation in *FLT3* gene.** Each data point was from a duplicate reaction and presented as individual blue dot. The error bar stands for one standard deviation. The fitted line is light blue. The Ct values and log VAFs exhibited liner correlation with an r-square is 0.996.

**(D). Standard VAF curve of 2503G>C (I836del) mutation in *FLT3* gene.** Each data point was from a duplicate reaction and presented as individual blue dot. The error bar stands for one standard deviation. The fitted line is light blue. The Ct values and log VAFs exhibited liner correlation with an r-square is 0.999.

**(E). Standard VAF curve of 388A>G (I130V) & 394C>T mutation in *IDH1* gene.** Each data point was from a duplicate reaction and presented as individual blue dot. The error bar stands for one standard deviation. The fitted line is light blue. The Ct values and log VAFs exhibited liner correlation with an r-square is 0.994.

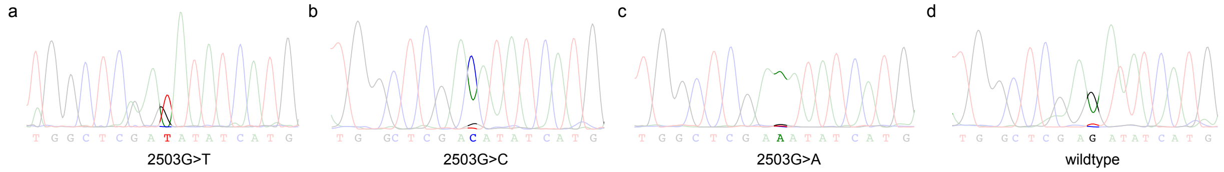


**Figure S11. Sanger Sequencing traces of 0.01% VAF gBlocks in *FLT3* gene.**

**(A). Sanger trace of 0.01% VAF *FLT3* 2503G>T synthetic reference sample.**

**(B). Sanger trace of 0.01% VAF *FLT3* 2503G>C synthetic reference sample.**

**(C). Sanger trace of 0.01% VAF *FLT3* 2503G>A synthetic reference sample.**

**(D). Sanger trace of wildtype gDNA sample.**

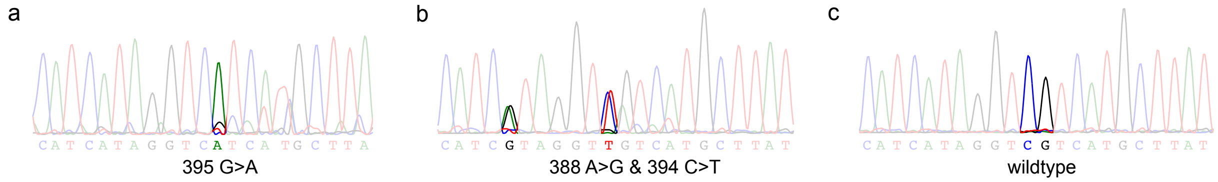


**Figure S12. Sanger Sequencing traces of 0.01% VAF gBlocks in *IDH1* gene.**

**(A). Sanger trace of 0.01% VAF *IDH1* 395G>A synthetic reference sample.**

**(B). Sanger trace of 0.01% VAF *IDH1* 388A>G & 394C>T synthetic reference sample.**

**(C). Sanger trace of wildtype gDNA sample.**

We further validated the 10% and 1% VAF samples of each gBlock variant in NGS. For each NGS library, 60ng sample in approximately 1.5 μL was input. Firstly, the same primer sets as in the Occlusion qPCR reactions and Phusion Hot Start Flex 2X Master Mix (Thermo Scientific) were used to amplify the samples; the thermocycling program started with 30 s polymerase activation and initial denaturation at 98 ˚C, followed by 25 cycles of 10 s at 98 ˚C for DNA denaturing, 30 s at 63 ˚C for annealing and 30 s at 72 ˚C for extension (98 ˚C: 30s - (98 ˚C: 10 s - 63 ˚C: 30 s - 72 ˚C: 30 s) x 25). Then a 2-cycle adaptor PCR step was used to append adaptor sequence to the amplicons, followed by TruSeq index PCR amplification. Then the concentrations of libraries were quantified by Qubit, the length of libraries was QCed in Bioanalyzer using high sensitivity DNA chips. Libraries were sequenced in 2 x 75 bp pair-end sequencing in Illumina Miseq instrument. The NGS result was summarized in Table S1.

| NGS | Var Reads | WT Reads | VAF |
|---|---|---|---|
| *IDH1* 395G>A 10% ref gBlock | 6,714 | 74,336 | 8.28% |
| *IDH1* 395G>A 1% ref gBlock | 747 | 76,542 | 0.97% |
| *IDH1* 388A>G & 394C>T 10% ref gBlock | 4,860 | 39,280 | 11.01% |
| *IDH1* 388A>G & 394C>T 1% ref gBlock | 558 | 49,788 | 1.11% |
| *FLT3* 2503G>T 10% ref gBlock | 21,038 | 275,557 | 7.09% |
| *FLT3* 2503G>T 1% ref gBlock | 2,528 | 288,950 | 0.87% |
| *FLT3* 2503G>C 10% ref gBlock | 18,914 | 277,118 | 6.39% |
| *FLT3* 2503G>C 1% ref gBlock | 2,609 | 285,145 | 0.91% |
| *FLT3* 2503G>A 10% ref gBlock | 23,561 | 278,276 | 7.81% |
| *FLT3* 2503G>A 1% ref gBlock | 2,932 | 363,980 | 0.80% |
| *FLT3* 2508_2510del 10% ref gBlock | 16,507 | 341,856 | 4.61% |
| *FLT3* 2508_2510del 1% ref gBlock | 1,605 | 382,071 | 0.42% |
| *DNMT3A* 2644C>T 10% ref gBlock | 8,748 | 103,754 | 7.78% |
| *DNMT3A* 2644C>T 1% ref gBlock | 770 | 85,557 | 0.89% |

**Table S1. Summary of VAF from NGS.**

**Section S5. The Occlusion qPCR and ddPCR results of PBMC samples from healthy donors and AML patients.**

27 PBMC samples from AML patients were purchased from discovery life science (DLS) and 15 PBMC samples from healthy donors were from the Zen-Bio. All the detailed sample information was shown in Table S2 and S3.

| Sample ID | Patient ID | Primary Diagnosis | Disease status | Gender | Patient Age at Collection | Draw Date | Treatment Status | Clinical Stage |
|---|---|---|---|---|---|---|---|---|
| 1 | 121713346 | Leukemia, Acute Myeloid (AML) | Stable | Male | 82 | 7/20/20 | Active | |
| 2 | 200000471 | Leukemia, Acute Myeloid (AML) | Stable | Male | 41 | 12/10/19 | Post | |
| 3 | 121592979 | Leukemia, Acute Myeloid (AML) | Stable | Male | 91 | 5/4/20 | Active | |
| 4 | 121582633 | Leukemia, Acute Myeloid (AML) | Stable | Female | 61 | 5/4/20 | Active | |
| 5 | 200003029 | MDS/AML | Stable | Male | 67 | 3/20/19 | Post | |
| 6 | 200008127 | Leukemia, Acute Myeloid (AML) | Newly Diagnosed | Female | 64 | 10/4/19 | Pre | M4 (FAB) |
| 7 | 200000471 | Leukemia, Acute Myeloid (AML) | Stable | Male | 41 | 12/10/19 | Post | |
| 8 | 110025045 | Leukemia, Acute Myeloid (AML) | Stable | Female | 61 | 7/7/17 | Post | M1 |
| 9 | 200009066 | Leukemia, Acute Myeloid (AML) | Newly Diagnosed | Male | Unknown | 7/14/20 | Active | |
| 10 | 200000340 | Leukemia, Acute Myeloid (AML) | Newly Diagnosed | Female | 38 | 5/15/19 | Pre | M4 |
| 11 | 200018300 | Leukemia, Acute Myeloid (AML) | Refractory | Male | 65 | 10/19/20 | Post | |
| 12 | 200009196 | Leukemia, Acute Myeloid (AML) | Stable | Female | 58 | 2/19/20 | Active | |
| 13 | 200013114 | Leukemia, Acute Myeloid (AML) | Unkown | Male | 83 | 10/29/19 | Pre | M5 |
| 14 | 200009609 | Leukemia, Acute Myeloid (AML) | Progressive | Female | 60 | 9/2/20 | Post | |
| 15 | 200018824 | Leukemia, Acute Myeloid (AML) | Newly Diagnosed | Female | 56 | 1/14/21 | Pre | M4 |
| 16 | 200018595 | Leukemia, Acute Myeloid (AML) | Relapse | Male | 56 | 12/7/20 | Post | M4 |
| 17 | 200009007 | Leukemia, Acute Myeloid (AML) | Refractory | Male | 53 | 2/25/20 | Post | M4 |
| 18 | 200003963 | Leukemia, Acute Myeloid (AML) | Refractory | Female | 55 | 12/17/19 | Active | |
| 19 | 200009536 | Leukemia, Acute Myeloid (AML) | Newly Diagnosed | Female | 61 | 5/4/20 | Pre | |
| 20 | 200009469 | Leukemia, Acute Myeloid (AML) | Progressive | Male | 71 | 12/8/20 | Post | |
| 21 | 200018487 | Leukemia, Acute Myeloid (AML) | Newly Diagnosed | Female | 49 | 12/22/20 | Pre | M5 |

| | | | | | | | | |
|---|---|---|---|---|---|---|---|---|
| 22 | 200009002 | Leukemia, Acute Myeloid (AML) | Newly Diagnosed | Female | 76 | 2/11/20 | Post | AML (Blast crisis after CML) |
| 24 | 200013554 | Leukemia, Acute Myeloid (AML) | Stable | Female | 70 | 12/11/19 | Post | |
| 25 | 200009189 | Leukemia, Acute Myeloid (AML) | Refractory | Female | 62 | 2/24/20 | Post | |
| 26 | 200005227 | Leukemia, Acute Myeloid (AML) | Unkown | Male | 62 | 9/10/19 | Post | M0 |
| 27 | 121624482 | Leukemia, Acute Myeloid (AML) | Stable | Female | 32 | 7/20/20 | Active | |
| 28 | 200018817 | Leukemia, Acute Myeloid (AML) | Relapse | Female | 64 | 12/18/20 | Post | |

**Table S2. Information of commercial AML samples.**

| Sample ID | Patient ID | Health Status | Quantity of Cells (M) / vial | Gender | Patient Age at Collection | BMI | Diabetic | Smoker |
|---|---|---|---|---|---|---|---|---|
| 25 | PBMC080621C | Healthy | 15 | Male | 58 | 25.8 | Unknown | Unknown |
| 26 | PBMC042121E | Healthy | 15 | Male | 72 | Unknown | Unknown | Unknown |
| 27 | PBMC063021C | Healthy | 15 | Male | 73 | Unkown | Unknown | Unknown |
| 28 | PBMC052621G | Healthy | 15 | Male | 55 | 29.9 | No | No |
| 29 | PBMC063021B | Healthy | 15 | Male | 27 | Unknown | Unknown | Unknown |
| 30 | PBMC080621B | Healthy | 15 | Female | 58 | 33.5 | Unknown | No |
| 31 | PBMC063021A | Healthy | 15 | Male | 75 | Unknown | Unknown | Unknown |
| 32 | PBMC090220C | Healthy | 15 | Male | 59 | Unknown | Unknown | Unknown |
| 33 | PBMC043019A | Healthy | 15 | Male | 64 | Unknown | Unknown | Unknown |
| 34 | PBMC051421F | Healthy | 15 | Male | 53 | 23.1 | Unknown | Yes |
| 35 | PBMC072121E | Healthy | 15 | Female | 65 | Unknown | Unknown | Unknown |
| 36 | PBMC052721A | Healthy | 15 | Male | 65 | 29.0 | No | Yes |
| 37 | PBMC090220E | Healthy | 15 | Male | 31 | Unknown | Unknown | Unknown |
| 38 | PBMC052721B | Healthy | 15 | Female | 19 | 24.6 | Unknown | No |
| 39 | PBMC090220G | Healthy | 15 | Male | 30 | Unknown | Unknown | Unknown |

**Table S3. Information of commercial healthy donors.**

We applied the BDA Occlusion qPCR reactions in 27 AML and 15 healthy PBMC samples purchased from the commercial vendors. Each sample was conducted in a duplicate reaction for each BDA Occlusion set, three BDA Occlusion sets in total. We used the *GAPDH* gene for quantification of the sample input, the obtained Ct values were summarized in Table S4 and S5. Then we input the same amount of the sample for *FLT3* and *IDH1* genes. All the collected Ct values were summarized in Table S6 to S9. The BDA Occlusion qPCR reported 6 AML patients with mutations in *FLT3* gene, 2 patients with mutations in *IDH1* gene (Figure S13). All the other samples from either AML patients or healthy donors were reported negative. PCR products of sample reported as mutant (positive) were sent for Sanger sequencing, which could further double confirm the genotype as well as identify detailed mutation information (Figure 4 and Figure S14). Knowing mutation information, we then called the VAF based on the fitting curves from the synthetic gBlock variants (Table S10).

We further validated our finding using ddPCR for *FLT3* gene, the results were summarized in Figure S15 to S18 and Table S11. The ddPCR data analysis using MATLAB was the same as previously described[1].

| *GAPDH* housekeeping gene | Ct value |
|---|---|
| AML #1 | 34.2 |
| AML #2 | 33.0 |
| AML #3 | 33.5 |
| AML #4 | 36.4 |
| AML #5 | 34.4 |
| AML #6 | 34.1 |
| AML #7 | 33.9 |
| AML #8 | 32.6 |
| AML #9 | 33.0 |
| AML #10 | 33.6 |
| AML #11 | 32.8 |
| AML #12 | 34.4 |
| AML #13 | 34.6 |
| AML #14 | 34.7 |
| AML #15 | 33.0 |
| AML #16 | 34.1 |
| AML #17 | 32.8 |
| AML #18 | 33.8 |

| | |
|---|---|
| AML #19 | 33.0 |
| AML #20 | 33.7 |
| AML #21 | 33.8 |
| AML #22 | 33.8 |
| AML #24 | 32.4 |
| AML #25 | 34.3 |
| AML #26 | 34.1 |
| AML #27 | 33.7 |
| AML #28 | 34.5 |
| gDNA NA18562 | 33.2 |

**Table S4. Ct value of AML samples using *GAPDH* BDA Occlusion set.**

| *GAPDH* housekeeping gene | Ct value |
|---|---|
| Healthy Donor #1 | 33.8 |
| Healthy Donor #2 | 34.6 |
| Healthy Donor #3 | 33.6 |
| Healthy Donor #4 | 34.0 |
| Healthy Donor #5 | 33.6 |
| Healthy Donor #6 | 34.1 |
| Healthy Donor #7 | 33.6 |
| Healthy Donor #8 | 34.2 |
| Healthy Donor #9 | 33.4 |
| Healthy Donor #10 | 34.1 |
| Healthy Donor #11 | 34.3 |
| Healthy Donor #12 | 33.9 |
| Healthy Donor #13 | 34.3 |
| Healthy Donor #14 | 33.2 |
| Healthy Donor #15 | 33.9 |

**Table S5. Ct value of healthy donors using *GAPDH* BDA Occlusion set.**

| *FLT3* gene | Ct value | Genotype |
|---|---|---|
| AML #1 | 43.2 | Wildtype |
| AML #2 | 43.1 | Wildtype |
| AML #3 | 41.5 | Wildtype |
| AML #4 | 42.8 | Wildtype |
| AML #5 | 42.5 | Wildtype |
| AML #6 | 43.1 | Wildtype |

| | | |
|---|---|---|
| AML #7 | 42.5 | Wildtype |
| AML #8 | 43.6 | Wildtype |
| AML #9 | 38.5 | Mutant |
| AML #10 | 36.3 | Mutant |
| AML #11 | 44.7 | Wildtype |
| AML #12 | 40.1 | Mutant |
| AML #13 | 43.9 | Wildtype |
| AML #14 | 27.2 | Mutant |
| AML #15 | 26.4 | Mutant |
| AML #16 | 43.3 | Wildtype |
| AML #17 | 43.7 | Wildtype |
| AML #18 | 41.2 | Wildtype |
| AML #19 | 42.4 | Wildtype |
| AML #20 | 47.9 | Wildtype |
| AML #21 | 44 | Wildtype |
| AML #22 | 41.8 | Wildtype |
| AML #24 | 32.2 | Mutant |
| AML #25 | 42.5 | Wildtype |
| AML #26 | 42.6 | Wildtype |
| AML #27 | 41.7 | Wildtype |
| AML #28 | 43 | Wildtype |
| gDNA NA18562 | 43.2 | Wildtype |

**Table S6. Ct value of AML samples using *FLT3* BDA Occlusion set**

| *FLT3* gene | Ct value | Genotype |
|---|---|---|
| Healthy Donor #1 | 42.7 | Wildtype |
| Healthy Donor #2 | 41.2 | Wildtype |
| Healthy Donor #3 | 42.9 | Wildtype |
| Healthy Donor #4 | 43.7 | Wildtype |
| Healthy Donor #5 | 42.1 | Wildtype |
| Healthy Donor #6 | 44.3 | Wildtype |
| Healthy Donor #7 | 42.0 | Wildtype |
| Healthy Donor #8 | 42.0 | Wildtype |
| Healthy Donor #9 | 41.6 | Wildtype |
| Healthy Donor #10 | 41.7 | Wildtype |
| Healthy Donor #11 | 43.4 | Wildtype |
| Healthy Donor #12 | 43.3 | Wildtype |
| Healthy Donor #13 | 41.6 | Wildtype |
| Healthy Donor #14 | 42.5 | Wildtype |

| | | |
|---|---|---|
| Healthy Donor #15 | 43.7 | Wildtype |

**Table S7. Ct value of healthy donors using *FLT3* BDA Occlusion set.**

| *IDH1* gene | Ct value | Genotype |
|---|---|---|
| AML #1 | 43.1 | Wildtype |
| AML #2 | 42.0 | Wildtype |
| AML #3 | 41.6 | Wildtype |
| AML #4 | 41.5 | Wildtype |
| AML #5 | 41.6 | Wildtype |
| AML #6 | 42.0 | Wildtype |
| AML #7 | 40.5 | Wildtype |
| AML #8 | 42.9 | Wildtype |
| AML #9 | 41.9 | Wildtype |
| AML #10 | 39.7 | Wildtype |
| AML #11 | 42.2 | Wildtype |
| AML #12 | 41.6 | Wildtype |
| AML #13 | 40.2 | Wildtype |
| AML #14 | 41.2 | Wildtype |
| AML #15 | 41.9 | Wildtype |
| AML #16 | 42.0 | Wildtype |
| AML #17 | 40.4 | Wildtype |
| AML #18 | 42.5 | Wildtype |
| AML #19 | 35.9 | Mutant |
| AML #20 | 39.8 | Wildtype |
| AML #21 | 40.8 | Wildtype |
| AML #22 | 41.3 | Wildtype |
| AML #24 | 27.7 | Mutant |
| AML #25 | 40.7 | Wildtype |
| AML #26 | 42.0 | Wildtype |
| AML #27 | 42.0 | Wildtype |
| AML #28 | 42.2 | Wildtype |
| gDNA NA18562 | 39.5 | Wildtype |

**Table S8. Ct value of AML samples using *IDH1* BDA Occlusion set.**

| *IDH1* gene | Ct value | Genotype |
|---|---|---|
| Healthy Donor #1 | 42.0 | Wildtype |
| Healthy Donor #2 | 41.8 | Wildtype |

| | | |
|---|---|---|
| Healthy Donor #3 | 41.5 | Wildtype |
| Healthy Donor #4 | 41.0 | Wildtype |
| Healthy Donor #5 | 43.9 | Wildtype |
| Healthy Donor #6 | 44.3 | Wildtype |
| Healthy Donor #7 | 41.0 | Wildtype |
| Healthy Donor #8 | 41.7 | Wildtype |
| Healthy Donor #9 | 39.5 | Wildtype |
| Healthy Donor #10 | 42.0 | Wildtype |
| Healthy Donor #11 | 42.8 | Wildtype |
| Healthy Donor #12 | 41.6 | Wildtype |
| Healthy Donor #13 | 42.5 | Wildtype |
| Healthy Donor #14 | 41.8 | Wildtype |
| Healthy Donor #15 | 41.4 | Wildtype |

**Table S9. Ct value of healthy donors using *IDH1* BDA Occlusion set**

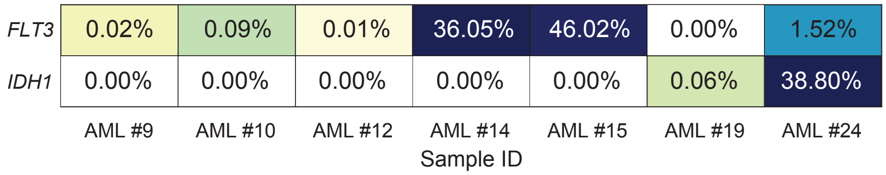


**Figure S13. Heatmap summary of AML samples reported positive in the *FLT3* or *IDH1* genes.**

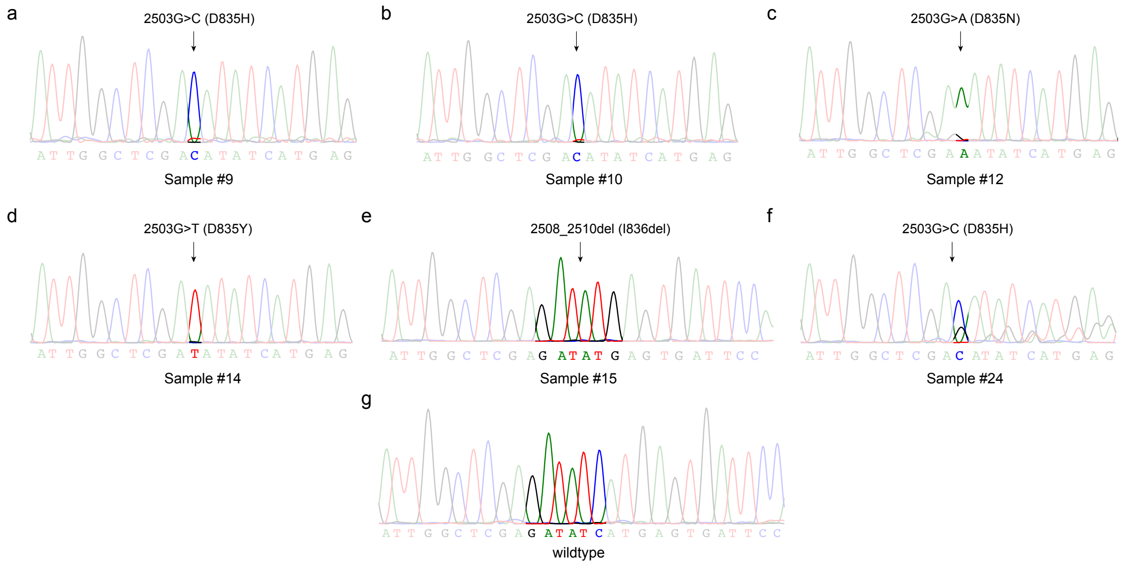


**Figure S14. Sanger traces of AML samples in *FLT3* gene**

| | Ct value | Mutation | Called VAF | Gene | Ct value of *GAPDH* set |
|---|---|---|---|---|---|
| AML sample #9 | 38.5 | 2503G>C | 0.02% | *FLT3* | 33.0 |
| AML sample #10 | 36.3 | 2503G>C | 0.09% | | 33.6 |
| AML sample #12 | 40.1 | 2503G>A | 0.01% | | 34.4 |
| AML sample #14 | 27.2 | 2503G>T | 36.05% | | 34.7 |
| AML sample #15 | 26.4 | 2508_2510del | 32.81% | | 33.0 |
| AML sample #24 | 32.2 | 2503G>C | 1.63% | | 32.4 |
| AML sample #19 | 35.9 | 395G>A | 0.06% | *IDH1* | 33.0 |
| AML sample #24 | 27.7 | 388A>G & 394C>T | 38.80% | | 32.4 |

**Table S10. Information summary of samples detected and called mutations in *FLT3* and *IDH1* genes**

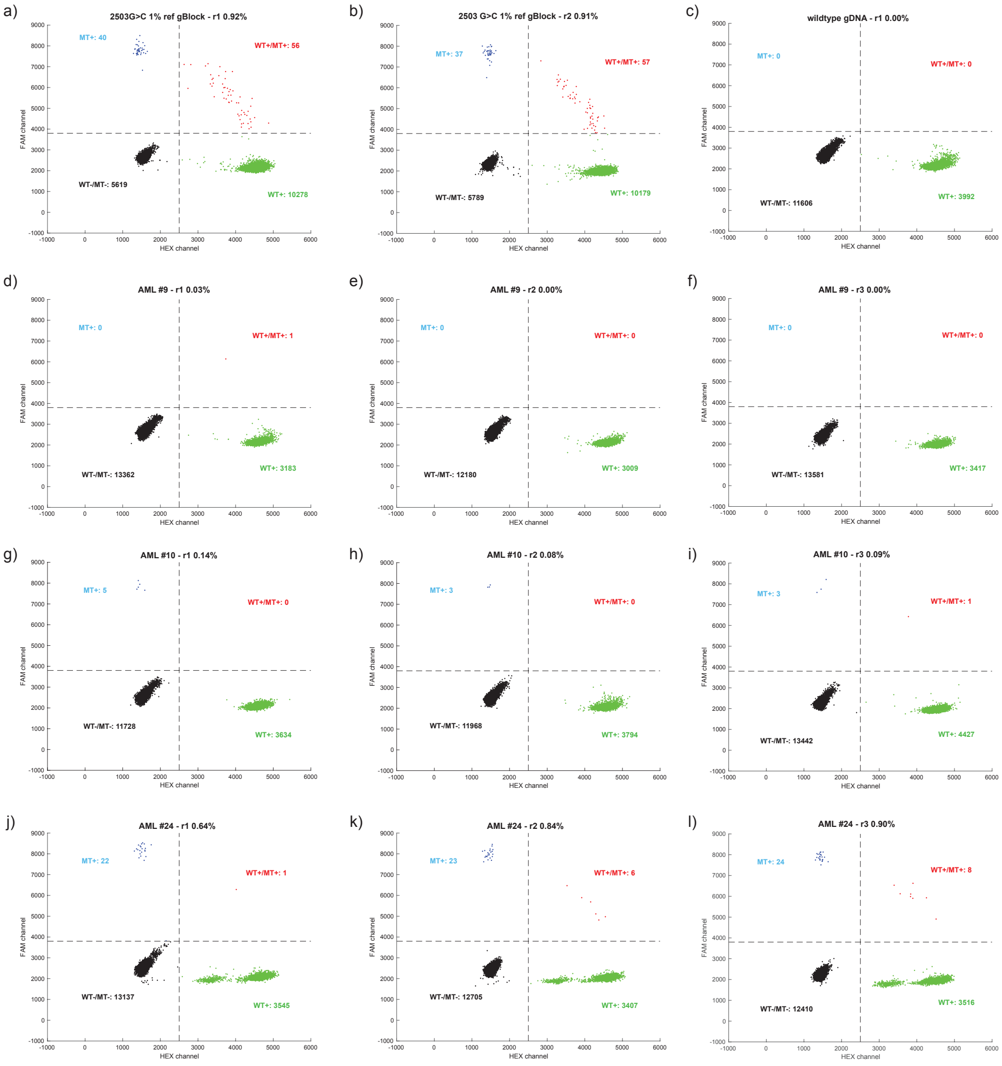


**Figure S15. ddPCR in *FLT3* 2503 G>C mutation**

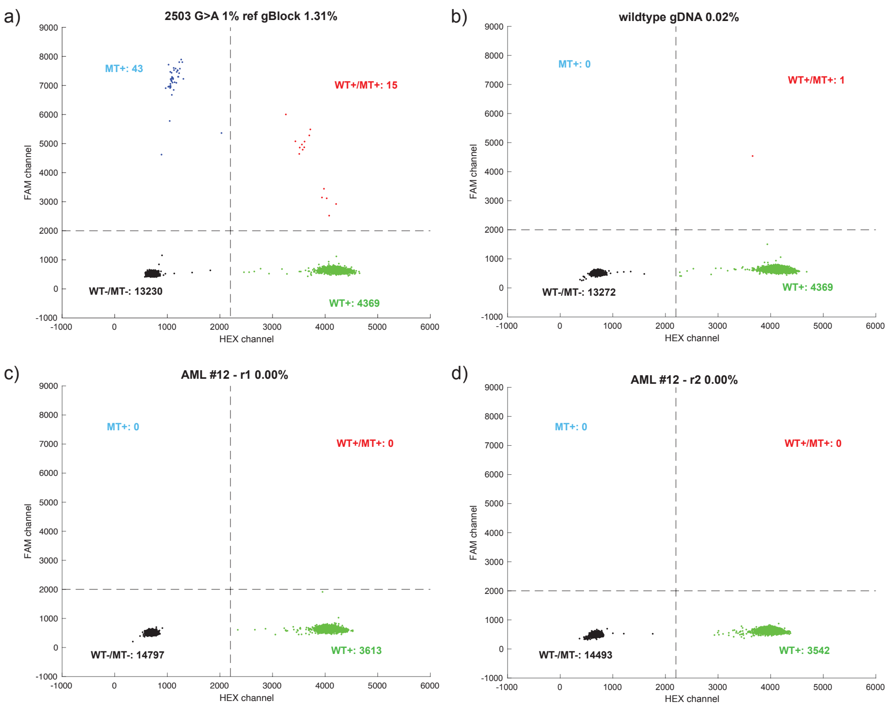


**Figure S16. ddPCR in *FLT3* 2503 G>A mutation**

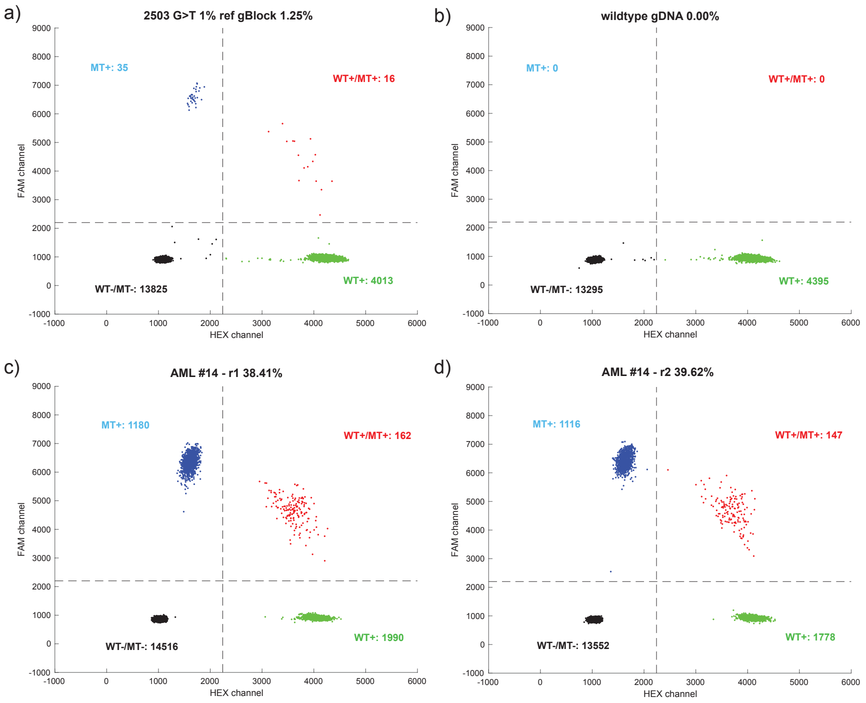


**Figure S17. ddPCR in *FLT3* 2503 G>T mutation**

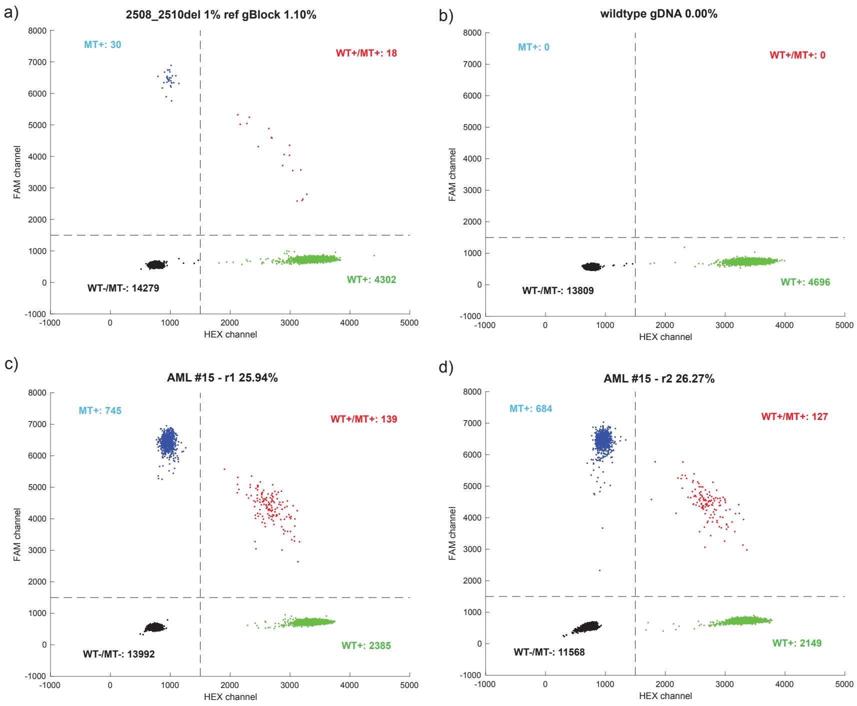


**Figure S18. ddPCR in *FLT3* 2508_2510del mutation**

| ddPCR | Var Counts | Wildtype Counts | VAF (%) | Mutation |
|---|---|---|---|---|
| 1% ref gBlock - r1 | 96 | 10334 | 0.92% | 2503 G>C |
| 1% ref gBlock - r2 | 94 | 10236 | 0.91% | |
| Wildtype gDNA | 0 | 3992 | 0.00% | |
| AML #9 - r1 | 1 | 3183 | 0.03% | |
| AML #9 - r2 | 0 | 3009 | 0.00% | |
| AML #9 - r3 | 0 | 3417 | 0.00% | |
| AML #10 - r1 | 5 | 3634 | 0.14% | |
| AML #10 - r2 | 3 | 3794 | 0.08% | |
| AML #10 - r3 | 4 | 4428 | 0.09% | |
| AML #24 - r1 | 23 | 3546 | 0.64% | |
| AML #24 - r2 | 29 | 3413 | 0.84% | |
| AML #24 - r3 | 32 | 3524 | 0.90% | |
| 1% ref gBlock | 58 | 4384 | 1.31% | 2503 G>A |
| Wildtype gDNA | 1 | 4640 | 0.02% | |
| AML #12 - r1 | 0 | 3613 | 0.00% | |
| AML #12 - r2 | 0 | 3542 | 0.00% | |
| 1% ref gBlock | 51 | 4029 | 1.25% | 2503 G>T |
| Wildtype gDNA | 0 | 4395 | 0.00% | |

| | | | | |
|---|---|---|---|---|
| AML #14 - r1 | 1342 | 2152 | 38.41% | |
| AML #14 - r2 | 1263 | 1925 | 39.62% | |
| 1% ref gBlock | 48 | 4320 | 1.10% | 2508_2510 del |
| Wildtype gDNA | 0 | 4696 | 0.00% | |
| AML #15 - r1 | 884 | 2524 | 25.94% | |
| AML #15 - r2 | 811 | 2276 | 26.27% | |

**Table S11. Summary of ddPCR results in mutations in *FLT3* gene.**

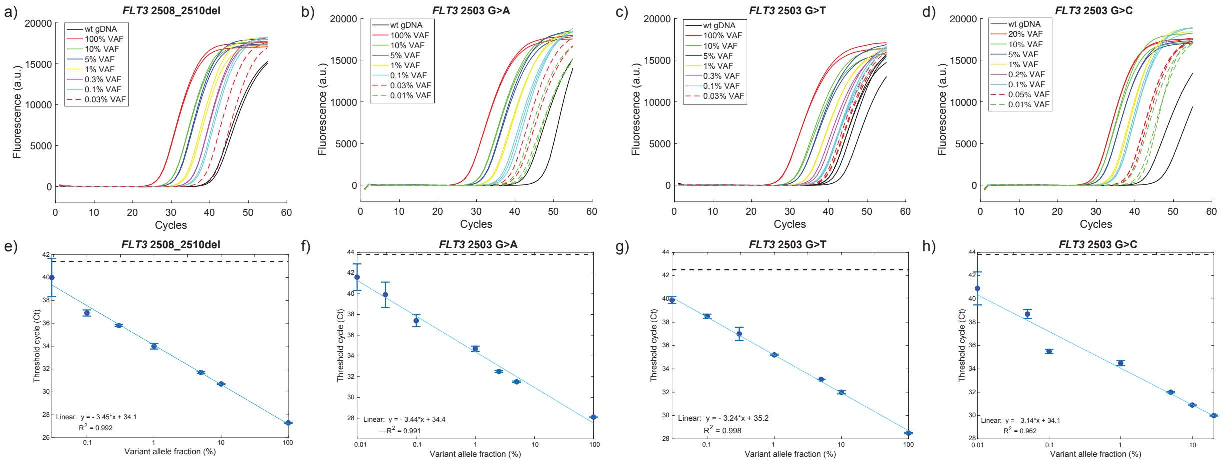


**Figure S19. Standard VAF curve of mutations.**

**(A-D). qPCR result for *FLT3* mutations.** Various mixtures of synthetic gBlock variants and genomic DNA with different VAFs were input, each mixture was performed in duplicate. For mutations 2508_2510del and 2503G>A, the amplification curves between 0.03% VAF (0.01% VAF for 2503G>A mutation) and the wildtype mutation were sometimes overlapped, this is suspicious of high variance in the number of input molecules within the low-VAF samples. For the 0.01% VAF sample in 30 ng input, the number of input molecules is roughly around one, which may be varied much in each reaction based on Poisson distribution.

**(E-H). Standard VAF curves of mutations in *FLT3* gene.** Each data point was from a duplicate reaction and presented as individual blue dot. The error bar stands for one standard deviation. The fitted line is light blue. The Ct values and log VAFs exhibited liner correlation with an r-square is 0.992 for 2508_2510del mutation, 0.991 for 2503G>A, 0.998 for 2503G>T and 0.962 for 2503G>C.

**Section S6. List of the Occlusion Probe component concentration and oligonucleotide sequences.**

Detailed information regarding the concentration of each component of each Occlusion qPCR reaction was provided in Table S12 to S14. All the sequences of oligonucleotides and synthetic gBlocks used were attached below (Table S15 and S16).

| Component | Stock concentration | Final concentration | Vol added (μL) |
|---|---|---|---|
| Phusion Hot Start Flex 2X Master Mix | 2X | 1X | 10 |
| Oligo Mix | 100 μM | 400 nM | 2 |
| DNA Template | 40 ng/μL | 200 ng | 5 |
| Water | | | 3 |

**Table S12. Detailed concentration of each component in Occlusion qPCR reaction.**

| Oligo Mix - BDA | Stock concentration | Final concentration | Vol added (μL) |
|---|---|---|---|
| Forward primer | 100 μM | 400 nM | 4.8 |
| Blocker | 100 μM | 4 μM | 48 |
| Occlusion primer | 100 μM | 400 nM | 4.8 |
| Occlusion probe | 100 μM | 400 nM | 4.8 |
| 0.1X TE buffer | | | 57.6 |

**Table S13. Formulation of oligo mix in Occlusion BDA qPCR reactions.**

| Oligo Mix | Stock concentration | Final concentration | Vol added (μL) |
|---|---|---|---|
| Forward primer | 100 μM | 400 nM | 4.8 |
| Occlusion primer | 100 μM | 400 nM | 4.8 |
| Occlusion probe | 100 μM | 400 nM | 4.8 |
| 0.1X TE buffer | | | 105.6 |

**Table S14. Formulation of oligo mix in a general Occlusion qPCR reaction.**

| | |
|---|---|
| Fig1_FP | TGATATGATTTTTAATTCCAAATGCTTAATGGA |
| Fig1_RP_Occlusion Primer-Cy5 | /5Cy5/CAAGCAATCAGGCGATCCGTTATGTaagagaacatgtttggttttgtgtaatg |
| Fig1_Occlusion Probe_LongStem | ACATAACGGAgCGCCTGAgTGCTTGGTGCTGTCACACTTGGTTACGCAAGTAACCAAGTGTGACAGCAC/3IAbRQSp/ |
| Fig1_Occlusion Probe_NoHairpin | ACATAACGGATCGCCTGATTGCTTG /3IAbRQSp/ |
| Fig1_Occlusion Probe_OptimalHairpin | ACATAACGGAgCGCCTGAgTGCTTGGTTCGCAAGAAC /3IAbRQSp/ |
| *FLT3*_FP | CCCCTGACAACATAGTTGGAATCA |

| | |
|---|---|
| *FLT3*_RP | GCTTGTCACCCACGGGAAA |
| *FLT3*_Occlusion Primer | /5Cy5/CAGGAAACAGCTATGACCGACAATGTGCTTGTCACCCACGGGAAA |
| *FLT3*_BDA blocker | ATAGTTGGAATCACTCATGATATCTCGAGCCA/iSpC3//iSpC3/CA |
| *FLT3*_Occlusion Probe | ACATTGTCGGgCATAGCTGgTTCCTGGTTCGCAAGAAC/3IAbRQSp/ |
| *DNMT3A*_FP | CAGCAGTCTCTGCCTCGC |
| *DNMT3A*_RP | CCTGCCCTCTCTGCCTTTT |
| *DNMT3A*_Occlusion Primer-HEX | /5HEX/TGTAAAACGACGGCCAGTACAAATGTCCTGCCCTCTCTGCCTTTT |
| *DNMT3A_BDA blocker* | CCTCGCCAAGCGGCTCATGTTG/iSpC3//iSpC3/GT |
| DNMT3A_Occlusion Probe-HEX/FAM | ACATTTGTACgGGCCGTCGgTTTACAGTTCGCAAGAAC/3IABkFQ/ |
| *DNMT3A*_Occlusion Primer-FAM | /56-FAM/TGTAAAACGACGGCCAGTACAAATGTCCTGCCCTCTCTGCCTTTT |
| *DNMT3A*_Occlusion Primer-ROX | /56-ROXN/TGTAAAACGACGGCCAGTACAAATGTCCTGCCCTCTCTGCCTTTT |
| DNMT3A_Occlusion Probe-ROX | ACATTTGTACgGGCCGTCGgTTTACAGTTCGCAAGAAC/3IAbRQSp/ |
| *GAPDH*_FP | CCTTCTTGCCTCTTGTCTCTTAG |
| *GAPDH*_RP | TCATTGATGGCAACAATATCCACT |
| *GAPDH*_Occlusion Primer | /56-ROXN/CAAGCAATCAGGCGATCCGTTATGTTCATTGATGGCAACAATATCCACT |
| *GAPDH*_BDA blocker | TTGTCTCTTAGATTTGGTCGTATTGGG/iSpC3//iSpC3/CA |
| *GAPDH*_Occlusion Probe | ACATAACGGAgCGCCTGAgTGCTTGGTTCGCAAGAAC/3IAbRQSp/ |
| *IDH1*_FP | CATGACTTACTTGATCCCCATAAGCA |
| *IDH1*_RP | AATCACCAAATGGCACCATACG |
| *IDH1*_Occlusion Primer | /5Cy5/CAAGCAATCAGGCGATCCGTTATGTAATCACCAAATGGCACCATACG |
| *IDH1*_BDA blocker | CCCATAAGCATGACGACCTATGATGATAGGTT/iSpC3//iSpC3/AA |
| *IDH1*_Occlusion Probe | ACATAACGGAgCGCCTGAgTGCTTGGTTCGCAAGAAC/3IAbRQSp/ |
| 80-plex-Forward-Occlusion Probe | CTGTCTCTTATACACATCTGACGCTGCCGACGAGTTCGCAAGAAC |
| 80-plex-Reverse-Occlusion Probe | CTGTCTCTTATACACATCTCCGAGCCCACGAGACGTTCGCAAGAAC |
| 179-plex-Forward-Occlusion Probe | TGATAGAgCGGAAGAGCGgCGTGTGTTCGCAAGAAC |
| 179-plex-Reverse-Occlusion Probe | AGAgCGGAAGAGCACACGgCTGTTCGCAAGAAC |

**Table S15. Sequences of oligonucleotides used.**

| | |
|---|---|
| FLT3_2503G>C<br>(D835H) | TCACTTTGTTTGTTGCACATCATCATGGCCGCTCACGGCACAGCCCAGTAAAGATAAGAGGCCTTCCATCACCGGTACCTCCTA<br>CTGAAGTTGAGTCTAGAAGAAAGATTGCACTCCAGGATAATACACATCACAGTAAATAACACTCTGGTGTCATTCTTGACAGTG<br>TGTTCACAGAGACCTGGCCGCCAGGAACGTGCTTGTCACCCACGGGAAAGTGGTGAAGATATGTGACTTTGGATTGGCTCGAcA<br>TATCATGAGTGATTCCAACTATGTTGTCAGGGGCAATGTGAGGCTGCTATTTCCTACTTATTTTTATACGGCTATTTTGTGTTG<br>TGTCGTTATCATGGTAAACAACTGCACTCACTGTGGTGCATTTTTGATTTATGGTAACATCAAAAAACCCTCACAGCAGTCTGC<br>TTACTTATGCTTAAAAGGTTTTTCTGCAGCTTCAGGGAATCTTCCATCTATAATAAAAGCTGAGCAAAATAGCATCACAT |
| FLT3_2503G>A<br>(D835N) | TCACTTTGTTTGTTGCACATCATCATGGCCGCTCACGGCACAGCCCAGTAAAGATAAGAGGCCTTCCATCACCGGTACCTCCTA<br>CTGAAGTTGAGTCTAGAAGAAAGATTGCACTCCAGGATAATACACATCACAGTAAATAACACTCTGGTGTCATTCTTGACAGTG<br>TGTTCACAGAGACCTGGCCGCCAGGAACGTGCTTGTCACCCACGGGAAAGTGGTGAAGATATGTGACTTTGGATTGGCTCGAaA<br>TATCATGAGTGATTCCAACTATGTTGTCAGGGGCAATGTGAGGCTGCTATTTCCTACTTATTTTTATACGGCTATTTTGTGTTG<br>TGTCGTTATCATGGTAAACAACTGCACTCACTGTGGTGCATTTTTGATTTATGGTAACATCAAAAAACCCTCACAGCAGTCTGC<br>TTACTTATGCTTAAAAGGTTTTTCTGCAGCTTCAGGGAATCTTCCATCTATAATAAAAGCTGAGCAAAATAGCATCACAT |
| FLT3_2503G>T<br>(D835Y) | TCACTTTGTTTGTTGCACATCATCATGGCCGCTCACGGCACAGCCCAGTAAAGATAAGAGGCCTTCCATCACCGGTACCTCCTA<br>CTGAAGTTGAGTCTAGAAGAAAGATTGCACTCCAGGATAATACACATCACAGTAAATAACACTCTGGTGTCATTCTTGACAGTG<br>TGTTCACAGAGACCTGGCCGCCAGGAACGTGCTTGTCACCCACGGGAAAGTGGTGAAGATATGTGACTTTGGATTGGCTCGAtA<br>TATCATGAGTGATTCCAACTATGTTGTCAGGGGCAATGTGAGGCTGCTATTTCCTACTTATTTTTATACGGCTATTTTGTGTTG<br>TGTCGTTATCATGGTAAACAACTGCACTCACTGTGGTGCATTTTTGATTTATGGTAACATCAAAAAACCCTCACAGCAGTCTGC<br>TTACTTATGCTTAAAAGGTTTTTCTGCAGCTTCAGGGAATCTTCCATCTATAATAAAAGCTGAGCAAAATAGCATCACAT |
| FLT3_2508_2510del<br>(I836del) | TCACTTTGTTTGTTGCACATCATCATGGCCGCTCACGGCACAGCCCAGTAAAGATAAGAGGCCTTCCATCACCGGTACCTCCTA<br>CTGAAGTTGAGTCTAGAAGAAAGATTGCACTCCAGGATAATACACATCACAGTAAATAACACTCTGGTGTCATTCTTGACAGTG<br>TGTTCACAGAGACCTGGCCGCCAGGAACGTGCTTGTCACCCACGGGAAAGTGGTGAAGATATGTGACTTTGGATTGGCTCGAGA<br>TATGAGTGATTCCAACTATGTTGTCAGGGGCAATGTGAGGCTGCTATTTCCTACTTATTTTTATACGGCTATTTTGTGTTGTGT<br>CGTTATCATGGTAAACAACTGCACTCACTGTGGTGCATTTTTGATTTATGGTAACATCAAAAAACCCTCACAGCAGTCTGCTTA<br>CTTATGCTTAAAAGGTTTTTCTGCAGCTTCAGGGAATCTTCCATCTATAATAAAAGCTGAGCAAAATAGCATCACAT |
| DNMT3A_2644C>T<br>(R882C) | TGATCTGAGTGCCGGGTTGTTTATAAAGGACAGAAGATTCGGCAGAACTAAGCAGGCGTCAGAGGAGTTGGTGGGTGTGAGTGC<br>CCCTGTCCCTGCACTTCGGGTGGCTGCTGGTCCTCCGGGTCCTGCTGTGTGGTTAGACGGCTTCCGGGCAGCCTGGTCTGGCCA<br>GCACTCACCCTGCCCTCTCTGCCTTTTCTCCCCCAGGGTATTTGGTTTCCCAGTCCACTATACTGACGTCTCCAACATGAGCtG<br>CTTGGCGAGGCAGAGACTGCTGGGCCGGTCATGGAGCGTGCCAGTCATCCGCCACCTCTTCGCTCCGCTGAAGGAGTATTTTGC<br>GTGTGTGTAAGGGACATGGGGGCAAACTGAGGTAGCGACACAAAGTTAAACAAACAAACAAAAAACACAAAACATAATAAAACA<br>CCAAGAACATGAGGATGGAGAGAAGTATCAGCACCCAGAAGAGAAAAAGGAATTTAAAACAAAAACCACAGAGGCGGAAA |
| IDH1_395G>A<br>(R132H) | TCGTGATGCCACCAACGACCAAGTCACCAAGGATGCTGCAGAAGCTATAAAGAAGCATAATGTTGGCGTCAAATGTGCCACTAT<br>CACTCCTGATGAGAAGAGGGTTGAGGAGTTCAAGTTGAAACAAATGTGGAAATCACCAAATGGCACCATACGAAATATTCTGGG<br>TGGCACGGTCTTCAGAGAAGCCATTATCTGCAAAAATATCCCCCGGCTTGTGAGTGGATGGGTAAAACCTATCATCATAGGTCa<br>TCATGCTTATGGGGATCAAGTAAGTCATGTTGGCAATAATGTGATTTTGCATGTTTTTTTTTTCATGGCCCAGAAATTTCCAAC<br>TTGTATGTGTTTTATTCTTATCTTTTGGTATCTACACCCATTAAGCAAGGTATGAAATTGAGAAATGCATATATGTATAACTGT<br>ATATTTACACACATTTAGCTAAAGGCAAATACAAATAAACTTACAAATAGGCGTCCATCTCAACACATTTTTTTTCAAAC |
| IDH1_388A>G&394C>T<br>(I130V&R132C) | TCGTGATGCCACCAACGACCAAGTCACCAAGGATGCTGCAGAAGCTATAAAGAAGCATAATGTTGGCGTCAAATGTGCCACTAT<br>CACTCCTGATGAGAAGAGGGTTGAGGAGTTCAAGTTGAAACAAATGTGGAAATCACCAAATGGCACCATACGAAATATTCTGGG<br>TGGCACGGTCTTCAGAGAAGCCATTATCTGCAAAAATATCCCCCGGCTTGTGAGTGGATGGGTAAAACCTATCATCgTAGGTtG<br>TCATGCTTATGGGGATCAAGTAAGTCATGTTGGCAATAATGTGATTTTGCATGTTTTTTTTTTCATGGCCCAGAAATTTCCAAC<br>TTGTATGTGTTTTATTCTTATCTTTTGGTATCTACACCCATTAAGCAAGGTATGAAATTGAGAAATGCATATATGTATAACTGT<br>ATATTTACACACATTTAGCTAAAGGCAAATACAAATAAACTTACAAATAGGCGTCCATCTCAACACATTTTTTTTCAAAC |

**Table S16. Sequences of synthetic gBlocks used.**
